# The Causal Artificial Intelligence Clinician for early hemodynamic management of septic shock in ICU

**DOI:** 10.64898/2026.07.06.26357375

**Authors:** Giovanni Angelotti, Laura Azzimonti, Maurizio Cecconi, Marco Zaffalon

## Abstract

**Rationale:** Standardizing fluid and vasopressor resuscitation in septic shock is challenging due to patient heterogeneity.

**Objectives:** To identify optimal intravenous fluid and vasopressor doses in first six hours of ICU admission in septic shock.

**Methods:** Graphical causal inference models grounded in expert clinical knowledge were applied to estimate heterogeneous treatment effects, limiting bias from spurious correlations. The model was trained on 1,706 MIMIC admissions and externally validated on 1,450 eICU admissions. The primary outcome was in-hospital survival; clinical improvement was the secondary outcome.

**Measurements and Main Results:** The cohort comprised 3,156 participants (age 65 years [IQR 53-75]; 42.8% female). Deviation from vasopressor recommendations was associated with failed clinical improvement (median OR 1.26, 95% CI 1.21-1.32), the most stable association across twelve sensitivity configurations, and with in-hospital mortality (median OR 1.16, 95% CI 1.10-1.24), which was null in five of twelve. Fluid deviations yielded 1.12 (1.08-1.18) and 1.02 (.96-1.07) respectively. These associations are confounded by indication by construction. External validation AUROCs were 0.72 (95% CI 0.67-0.76) for survival and 0.70 (95% CI 0.66-0.73) for improvement, comparable to predictive baselines. Treatment response varied with baseline physiology, fluids showing neutral or positive effects and high-dose norepinephrine worse ones.

**Conclusions:** An expert-grounded causal framework produced recommendations that behaved as an informative policy in external validation, using one-third the variables of a conventional predictive baseline. Whether alignment with those recommendations improves outcomes requires prospective evaluation. Code and instructions to reproduce this analysis, including a dashboard to explore our results, are available at https://github.com/IDSIA/causal-ai-clinician.

**Impact statement:** This study shows that a causal model built from an explicit, clinician-derived graph can generate early fluid and vasopressor dose recommendations for septic shock while using approximately one-third of the variables required by a conventional predictive model. Because the assumptions behind each recommendation are stated in the causal graph rather than embedded in a black-box predictor, they can be inspected, contested and revised by clinicians. The dose recommendations themselves remain hypothesis-generating and require prospective evaluation before any clinical use.

## Introduction

The early hours for septic shock patients, which include the historically termed *golden hour*, represent a period of rapidly evolving tissue hypoperfusion and cytopathic hypoxia where delayed intervention causes irreversible organ dysfunction.^1,2,3^ Current clinical guidelines advocate for immediate fluid resuscitation and early vasopressor initiation to restore mean arterial pressure and optimize oxygen delivery.^4,5,6^ However, host-response heterogeneity frequently renders standardized protocols insufficient.^7,8^ Evidence suggests that tailoring the dosing of vasopressors and fluids in these first hours may play a key role in mitigating immediate hemodynamic instability and ultimately increasing the chances of survival.^9^ Determining the optimal treatment combination remains, however, challenging. The design of Randomized Controlled Trials (RCTs) is complicated by the extreme heterogeneity of sepsis, the lack of known biomarkers, and poorly understood dosing thresholds.^10,11^ Concurrently, intensive care units are among the most data-rich environments across clinical practice, offering vast repositories of observational data.^12,13^ Consequently, estimating the effect of resuscitation strategies in sepsis using these observational records has been a recurrent objective of modern artificial intelligence systems.^14,15^ These applications rely on predictive models that exploit statistical correlations to generate risk estimates for outcomes. However, they do not assist clinicians in identifying optimal treatment strategies to transition patient states toward improved clinical conditions.^16^ There is therefore a growing need for AI systems capable of operationalizing evidence as a bridge that *concretely* connects actions to patients outcomes.^17^ Causal inference offers a principled response to this need, providing the methodological tools to move beyond association and formally reason about the effects of interventions.^18^ These methods have an established history in healthcare, now gaining renewed momentum through the scalability of novel Machine Learning (ML) models,^19,20^ known as Causal AI.^21^

This manuscript adopts Structural Causal Models (SCMs), which represent causal relations as arcs in a graph.^22^ Predictions made by models built in this manner are guaranteed to be consistent with the encoded assumptions, thereby enabling the estimation of treatment effects under the appropriate conditions.^23^

In intensive care settings, SCMs have been applied to model oxygen therapy effects on mortality,^24^ health equity disparities,^25^ discharge decisions,^26^ and have been used to audit sepsis treatments models.^27^ Here, we introduce the *Causal Artificial Intelligence Clinician*, a framework that leverages SCMs to identify the optimal resuscitation strategy in the first six hours of intensive care for septic shock patients, formally encoding the clinical, physiological, and operational knowledge of healthcare professionals into a Causal AI model. In external validation the recommendations behaved as an informative policy should, deviation from them being associated with worse outcomes, most consistently for vasopressors and clinical improvement; this comparison cannot, however, be separated from confounding by indication. The framework remained robust to distribution shift while using fewer, more causally meaningful variables than predictive baselines, showing how encoding domain knowledge supports the estimation of treatment effects from the secondary use of electronic health records.

This analysis is structured into three main phases, as illustrated in Figure 1. First, an initial deliberation phase is conducted to identify a set of clinically robust assumptions (panel (a)). Then, the assumptions encoded in a causal graph guide the training of ML models (panel (b)). Finally, the resulting models are evaluated on an external validation set (panel (c)). TRIPOD+AI and Target Statement checklists for this study are reported in Supplementary D. Our work is fully reproducible, instructions to build the datasets and re-run our analysis is available at: https://github.com/IDSIA/causal-ai-clinician.

**Figure 1:**
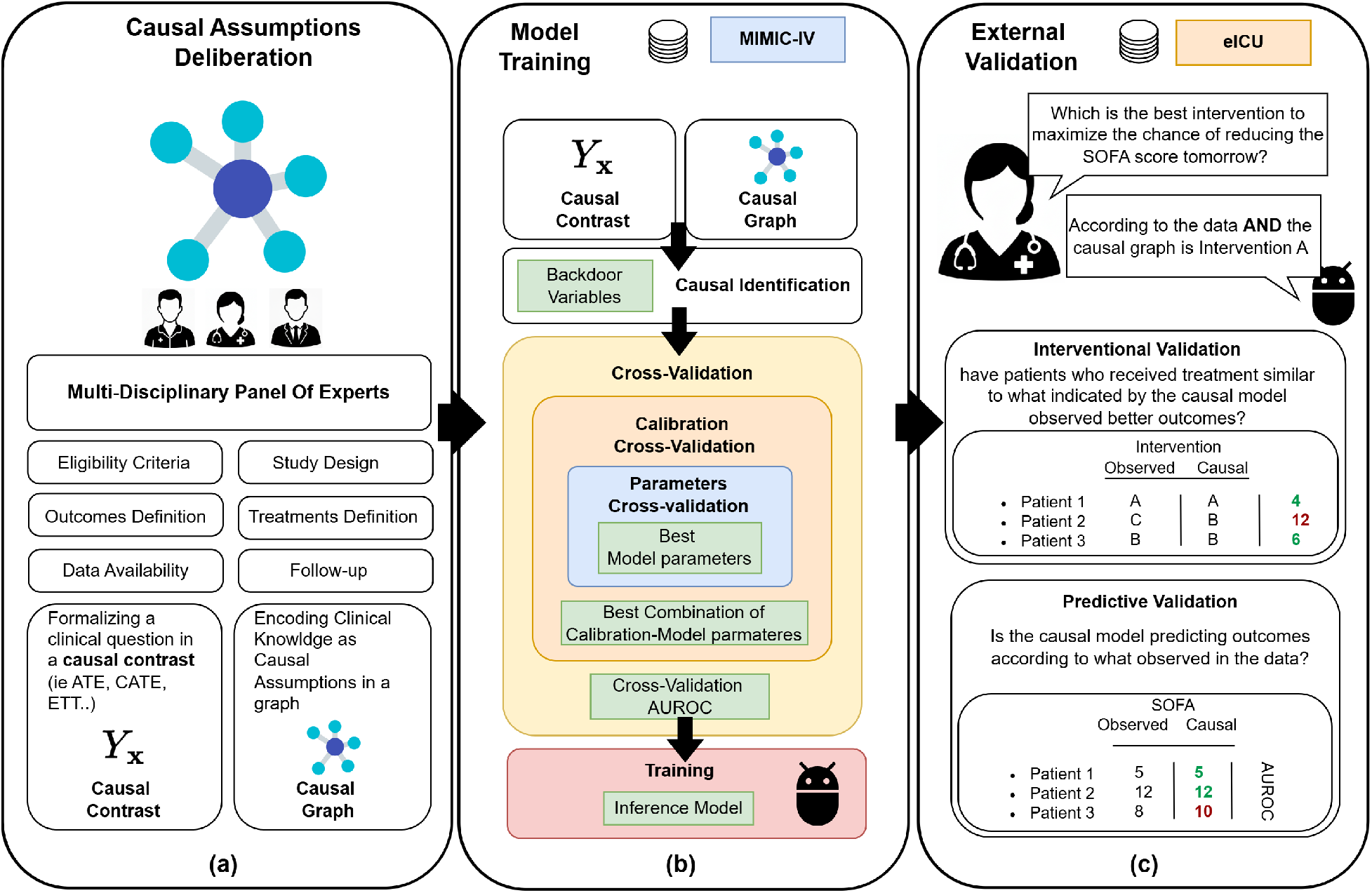
Graphical Abstract. Alt text: Three-panel schematic of the study workflow: a deliberation phase in which clinicians and methodologists agree a causal graph, a training phase in which the graph determines the feature set for the machine learning models, and an evaluation phase on the external validation cohort.

## Methods

### Assumptions and Causal Implications

This study employs SCMs to estimate the effect of intravenous fluid and vasopressor doses on patient outcomes from observational data, encoding clinical evidence and domain knowledge in a causal graph to separate causal effects from spurious correlations. This supports simulated interventions that quantify survival and improvement probabilities under specific treatment scenarios, a formal approximation of an RCT.

Specifically, for each patient, the model estimates the expected change in the probability of in-hospital survival, our primary endpoint, or clinical improvement, secondary outcome, when administering specific doses of norepinephrine and intravenous fluids compared to no treatment. This can be formalized as a *causal contrast*, here the Conditional Average Treatment Effect (CATE), as

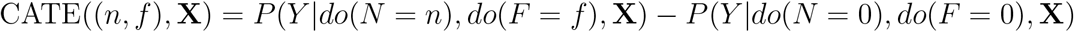

where the *do* operator fixes the fluid dose *F* and the norepinephrine dose *N* to the values *f* and *n* regardless of their causal parents, and *Y* = 1 denotes in-hospital survival or clinical improvement in a population with baseline clinical profile **X** (Supplementary C).

Estimating CATE via predictive models without causal adjustment can be biased, as it may incorrectly condition on descendants of the treatment.

We instead exploit the clinical knowledge encoded in the SCM to robustly measure the CATE from the data. We employ the *Backdoor Criterion* to identify which features should or should not be included in our model in order to properly recover the treatment effect. The Backdoor criterion can be verified from a causal graph alone and guarantees that if we can adjust for all parents **Z** of *F* and *N* the treatment effect can be recovered from the data^28^(for a rigorous formulation of the Backdoor definition, please refer to corollary 3 in^22^). If the criterion holds, we can convert the causal contrast into a statistical estimator solvable with common machine learning models:

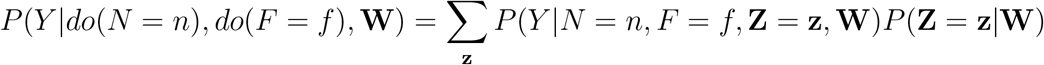

where **W** are the clinical descriptors in **X** that are not in **Z** and are non-descendants of *N* and *F*. Here we estimate the CATE using gradient boosting trees.We introduce a weak positivity assumption for all combinations of treatments within a clinically reasonable range and a smoothness assumption for the CATE surface. That is, positivity is limited by sample size rather than strictly violated, and similar treatments produce similar effects. Finally, we assume a static treatment regimen as estimating dynamic titration would require ad-hoc sequential methods which are not the scope of this investigation.

Details are provided in Supplementary C.

### Causal Graph Derivation

The causal graph was developed through iterative multidisciplinary consensus among clinicians and causal methodologists, encoding clinical assumptions about early hemodynamic management in septic shock. It incorporates both observed variables, retrieved from MIMIC and eICU, and unmeasured confounders not available in the Electronic Health Record (EHR). Further details on the derivation, and the full feature set, are given in Supplementary C and Table S1.

### Cohort Derivation

Data for this study were extracted from the MIMIC-IV v3.1 Database (MIMIC) and eICU v2.0 Collaborative Research Database (eICU). The MIMIC database contains EHR from ICU patients collected at the Beth Israel Deaconess Medical Center in Boston, US, from 2008 to 2022.^12^ The eICU is a multi-center EHR database with over 200,000 ICU admissions across the United States between 2014 and 2015.^13^ To isolate the septic patients of interest, we first restricted our cohort to all patients admitted to the hospital from the Emergency Department (ED). To ensure we captured the earliest available septic shock dynamics, we included only ICU admissions (T_0H_) occurring within 6 hours of the earliest available patient-hospital interaction (T_-6H_), defined as ED admission in MIMIC and hospital admission in eICU. We further restricted the minimum ICU stay length to 30 hours (T_30H_), the shortest time interval over which we could compute a complete updated SOFA score. This was necessary to compute our endpoint of clinical improvement, defined as the different between the worst 24-hour SOFA recorded in the [T30H,T54H] interval and the admission SOFA, estimated using data in the [T0H,T6H] window. The first 6 hours of the ICU stay [T_0H_, T_6H_] represent the window used for feature extraction.

We excluded patients admitted for surgery to avoid confounding due to trauma and patients with a Do Not Resuscitate (DNR) order during the first 30 hours of their ICU stay to ensure continuity of care. In eICU, we additionally excluded patients who were subsequently transferred to avoid survival bias induced by severe cases being transferred to other centers. Variables extracted from the first 6 hours of the ICU stay [T_0H_,T_6H_], were also used to filter our cohort. To reduce false-positive inclusions we applied three separate exclusions, each reported with its own count in Figure S2: approximate lactate below 2 mmol/L; no evidence of fluid or antibiotic therapy; and mean blood pressure of at least 65 mmHg with no evidence of vasopressor therapy. Norepinephrine equivalent dose was estimated by combining dopamine, epinephrine, norepinephrine, phenylephrine, and vasopressin based on Goradia et al. and Johnson et al.’s implementation.^29,30^ Norepinephrine equivalent dose was not documented for 29.6% of MIMIC and 77.5% of eICU admissions (Table S3); an undocumented dose was taken as zero, consistent with the absence of a recorded infusion. Details regarding the study design are shown in Figure S1 in Supplementary B.

### Model Training and Predictive Performance Assessment

We trained the causal model on the MIMIC dataset and externally validated it using eICU. The causal model used as input features the sufficient adjustment set defined by our causal graph (Figure 2), and was fitted with LightGBM gradient boosted trees. We used doubly nested cross-validation, applying an inner loop for hyperparameter tuning and an outer loop for calibration to accurately estimate treatment effects. Overall performance and fairness were evaluated via Area Under the Receiver Operating Characteristic curve (AUROC) over 2,000 bootstraps, stratifying by gender and race to assess parity against male and White majority reference groups. Further details are provided in Supplementary C.

**Figure 2:**
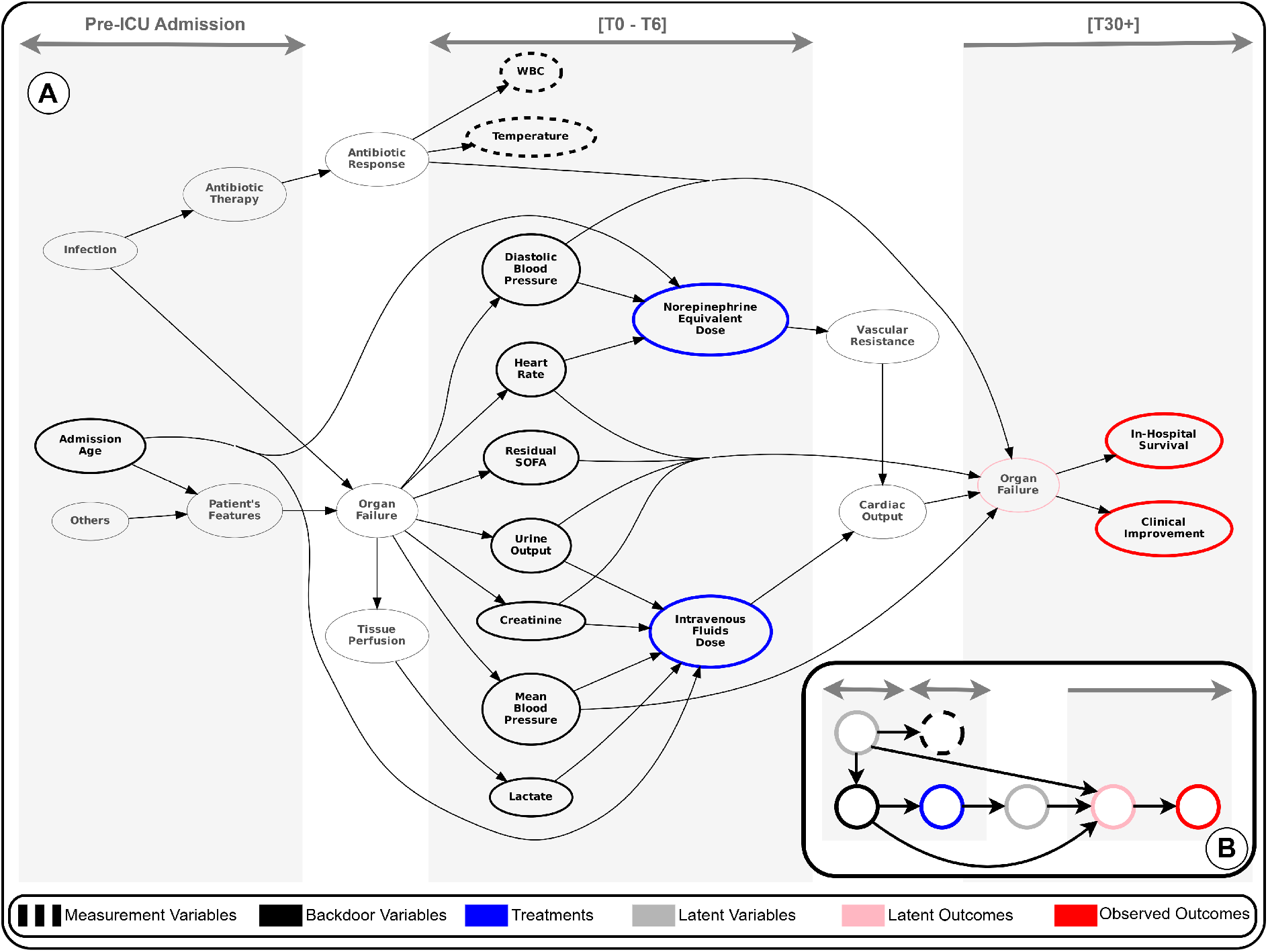
Causal Graph: DAG encoding the assumptions of our study. Outcomes are shown in red, decision variables in blue, backdoor variables in black, noisy proxies in dashed black, unmeasured variables in grey and unmeasured outcomes in pink. Figure shows a detailed (A) and exemplified (B) representation of the causal graph. Alt text: Directed acyclic graph of the study assumptions, shown in a detailed version and a simplified version. Outcomes are red, the two treatment decision variables blue, backdoor adjustment variables black, noisy proxies dashed black, unmeasured variables grey and unmeasured outcomes pink. Arrows run left to right from infection and antibiotic response, through organ failure and the physiological triggers of dosing, to the treatments and outcomes.

### Optimal Intervention Estimation and Validation

For each patient, the optimal combination of vasopressor and fluid doses was identified as the one maximizing the estimated CATE, evaluated across a discrete grid of clinically observed dose ranges. As a face-validity check rather than a causal test, patient outcomes in the external validation set were compared according to the deviation between administered treatments and model recommendations: an informative policy should show worse outcomes as deviation grows. Because clinicians titrate to severity, this comparison is confounded by indication by construction, and we therefore stratified it by SOFA. The association between treatment deviation and patient outcomes was quantified using repeated cross-validation and bootstrapping. Treatment effect heterogeneity across patient subgroups was further assessed by estimating CATEs conditioned on different baseline clinical profiles, with profiles clustered by common treatment-response trends to identify clinically meaningful examples. Further details are provided in Supplementary C.

### Sensitivity Analysis

We performed twelve sensitivity analyses to test design robustness across various inclusion and exclusion criteria, modeling choices, and missing data assumptions. Evaluations tested alternative feature sets, DNR inclusion, a shorter 24-hour window, treatment frequency thresholds, missing treatment exclusions, temporal alignment between databases, and staggered backdoor-treatment measurement windows; the twelve configurations are listed in full in Table S8. For each sensitivity analysis we measured how deviations from the optimal treatment, according to the causal model, correlated with outcomes. To quantify effects linked to confounding by indication, we measure strength of association for different severity strata according to patients’ SOFA score. We classified patients severity in two tiers: low (SOFA *<* 8) and high (SOFA *≥* 8).

## Results

### Study Design and Cohort Characterization

The study included 3,156 eligible admissions, utilizing MIMIC-IV for training and eICU for external validation. The primary endpoint was in-hospital survival and the secondary endpoint was clinical improvement, defined as a minimum two-point reduction in the SOFA within the 24 hours following the feature extraction window. These endpoints were observed in 81.1% and 12.2% of admissions, respectively. The analyzed treatments included total intravenous fluid intake and average norepinephrine equivalent dose during this initial window. See tables S1, S2 and S3 in Supplementary B for more details.

### Causal graph derivation and physiological grounding

We reconstructed the clinical reasoning process underlying the physician’s decision-making. At its core, the physician acts on two therapeutic interventions, fluid administration and norepinephrine, both aimed at modulating the downstream hemodynamic state: fluids influence cardiac output through preload optimization, while norepinephrine acts on systemic vascular resistance. We further assume that physicians rely on a set of specific physiological triggers to reach their treatment decisions. Fluid titration is driven by markers of hypoperfusion and renal stress, namely lactate, mean arterial pressure, and urine output. Norepinephrine doses, instead, are adjusted in response to diastolic pressure and heart rate, which themselves reflect the current degree of organ failure. Underlying this entire process is the complex interaction between the host’s response to infection, for which we assume that an appropriate antibiotic was selected earlier in the emergency room, and the patient’s age, which guides dose adjustment. Taken together, these assumptions are encoded in the causal graph shown in Figure 2. This graph allows us to translate our clinical question, namely:

> “What is the probability of survival or clinical improvement if we administer a different dosage combination of norepinephrine and intravenous fluid in the early hours following ICU admission in septic shock patients?”,

into a well-defined and rigorous causal contrast.

Furthermore, given that all treatments’ parents (black nodes, Figure 2) are adjusted for in our model, the backdoor criterion is verified, even in the presence of unmeasured confounding (grey nodes, infection, antibiotic therapy and antibiotic response).

### Optimal treatments, confounding and clinical outcomes

Deviation from the recommended vasopressor dose was most consistently associated with failed clinical improvement: each 1-SD increase in quantile-normalized deviation multiplied the odds of failing to improve by 1.26 (95% CI: [1.21, 1.32]), with restrictive fluids showing the same direction at 1.12 (95% CI: [1.08, 1.18]) (Figure 4b). The picture for in-hospital survival was weaker: vasopressor deviation multiplied the odds of death by 1.16 (95% CI: [1.10, 1.24]), while fluid deviation showed no association (1.02, 95% CI: [0.96, 1.07]) (Figure 4a, Table S8). These are associations between the delivered dose and the observed outcome, not estimates of the effect of following the policy. Non-linear mappings populate the dose-response space (Figure 4).

The model demonstrates predictive robustness, achieving external validation median AUROCs of 0.72 (95% CI: [0.67, 0.76]) for survival and 0.70 (95% CI: [0.66, 0.73]) for clinical improvement. This matches a gradient boosting baseline trained on a feature superset (Table S1; AUROCs: 0.74, 95% CI: [0.70, 0.78] and 0.68, 95% CI: [0.65, 0.72]; Figure 3, Table S5). Sociodemographic subgroup analyses were underpowered; the only formally significant gap was for clinical improvement in female patients (ΔAUROC *−*0.07, 95% CI: [*−*0.14, 0.00]), while performance in Hispanic patients was lower but imprecisely estimated (Table S4; calibration details in B.2).

**Figure 3:**
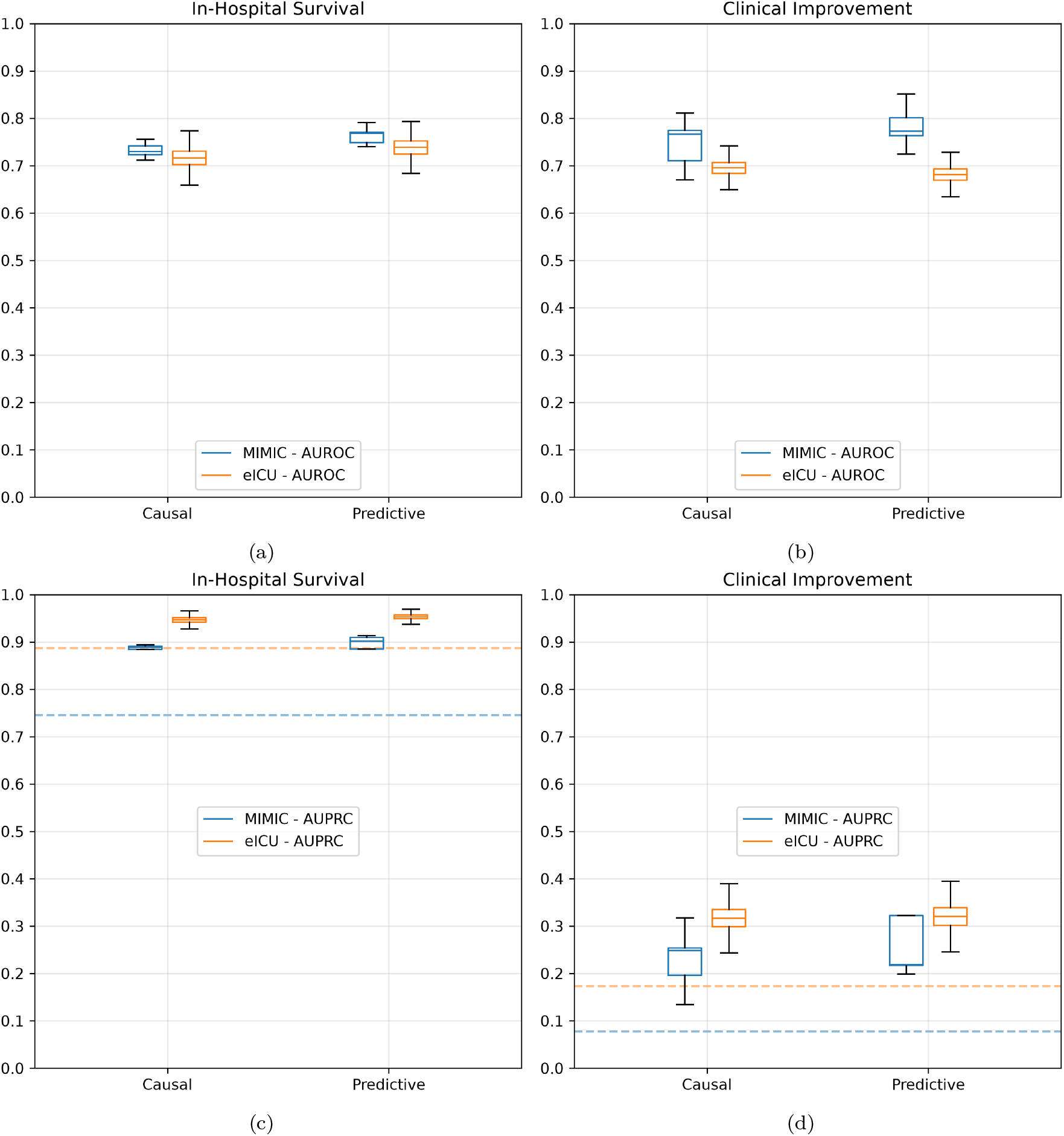
Causal and predictive model prediction performance the two study outcomes: in-hospital survival (a, c) and clinical improvement (b, d). Boxplots for AUROC (a, b) and AUPRC (c, d) evaluated in cross-validation on MIMIC (Blue) and in external validation on eICU (Orange). Dashed lines represent baseline prevalence. Alt text: Four box plots comparing the causal and predictive models. The top row shows AUROC and the bottom row AUPRC, with in-hospital survival on the left and clinical improvement on the right. Blue boxes are cross-validation on MIMIC and orange boxes external validation on eICU; dashed horizontal lines mark outcome prevalence.

**Figure 4:**
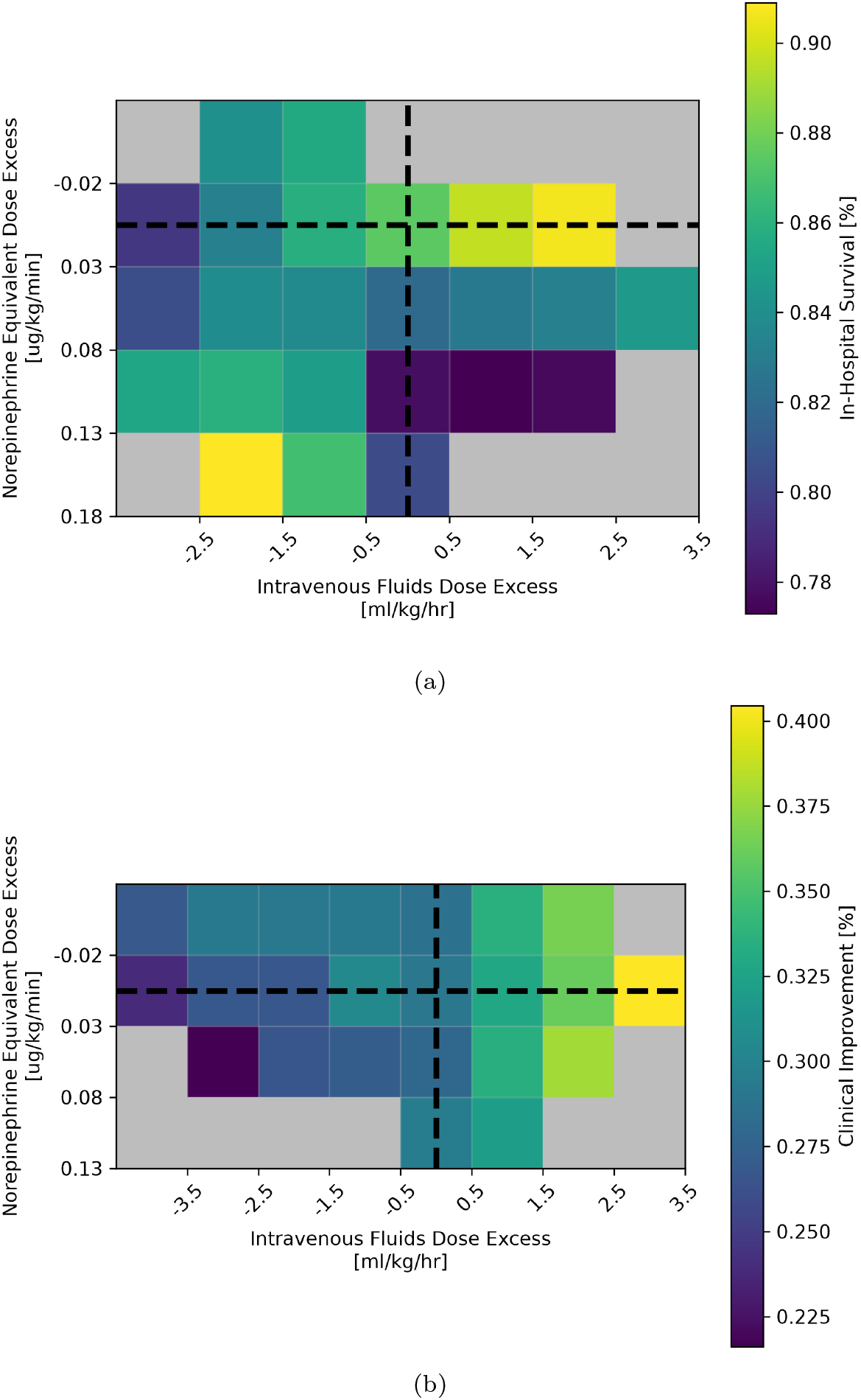
External interventional validation: heatmaps representing outcomes prevalence in relation to the difference between observed prescribed dose in eICU and predicted optimal dose according to the causal model for (a) in-hospital survival and (b) clinical improvement. Lighter shades indicate groups of patients with a higher prevalence of better outcomes, while darker shades indicate poorer outcomes. The horizontal black dotted line represents optimal norepinephrine equivalent dose, the vertical dotted line represents optimal intravenous fluid dose. Their intersection represents the optimal combination of intervenable variables. Alt text: Two heatmaps of outcome prevalence in eICU against the difference between the delivered dose and the dose recommended by the causal model, for in-hospital survival and for clinical improvement. The horizontal axis is fluid deviation and the vertical axis norepinephrine deviation. Lighter shading marks better outcomes, darker shading worse. Dotted lines mark the recommended dose of each treatment and their intersection the recommended combination.

### Heterogeneity of the treatment effect

Investigating dose-response profiles conditioned on baseline physiologies reveals significant heterogeneity across patient subgroups (Figure 5). Intravenous fluid administration generally yields neutral or positive median treatment effects, though patient s130 exhibits initial negative effects before benefiting at higher doses. Conversely, norepinephrine escalation frequently produces negative treatment effects, indicating probable harm at elevated doses. Confidence intervals widen considerably at higher norepinephrine doses (profiles s157 and s56) and, despite stable median predictions, remain wide for fluids in patient s14. Patients exhibiting declining prognoses with escalating vasopressors, such as s157, present with elevated baseline heart rates (109.7 BPM) and low mean blood pressures (59.3 mmHg) indicating severe physiological stress. Profiles showing stable responses to fluids, like s38, exhibit lower severity scores, including a 0.0 residual SOFA and lower lactate (2.2 mmol/L). These variations emphasize that therapeutic efficacy varies with individual baseline hemodynamic and metabolic variables. See Table S11 in Supplementary C for a complete clinical characterization of these examples.

**Figure 5:**
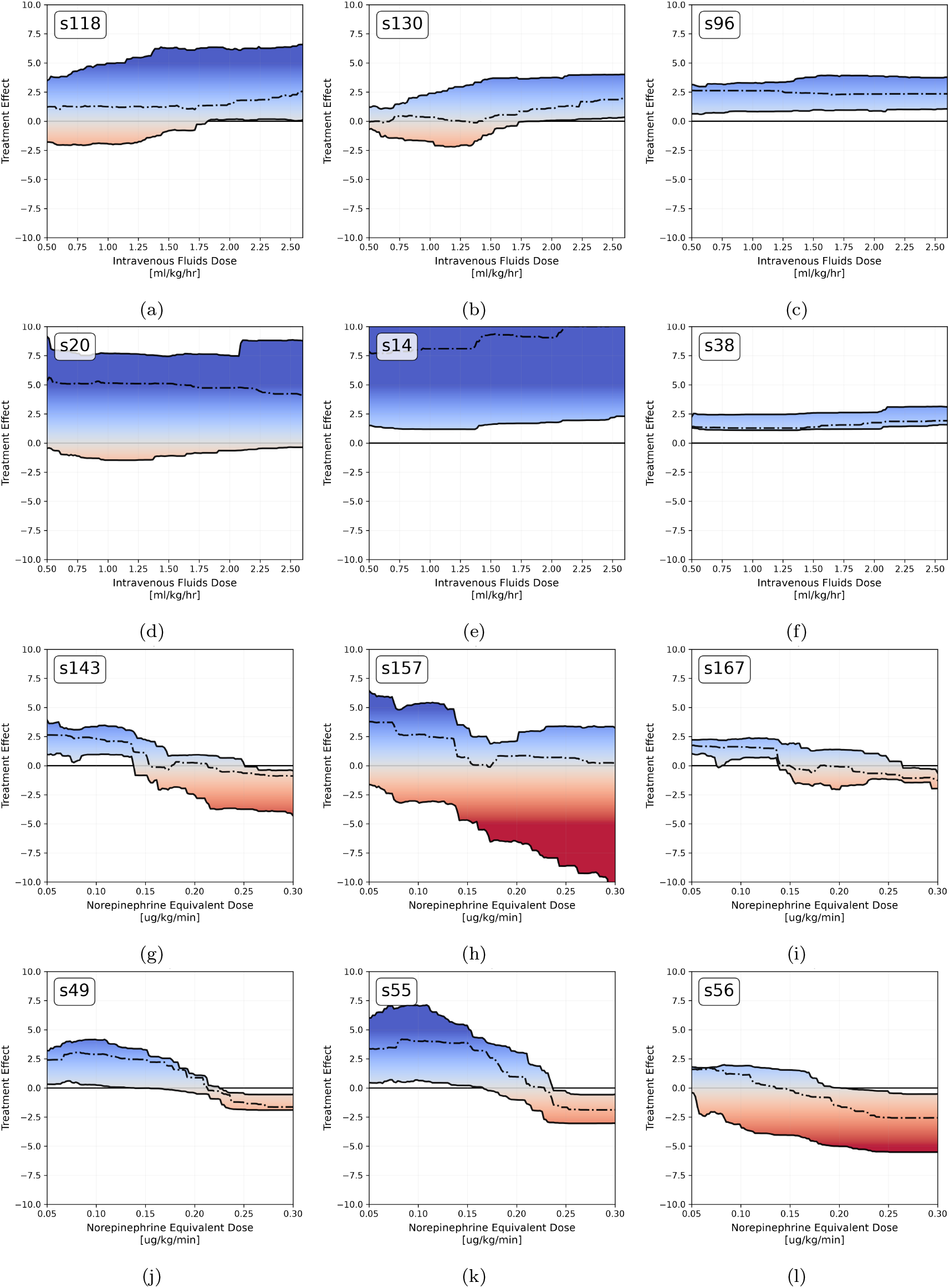
Simulated interventions: change in the outcome probability predicted by the causal model, in percentage points (y-axis) as a function of simulated dose (x-axis). Positive values indicate a shift toward better prognosis, negative values a harmful treatment effect. Panels (a-c) and (d-f) show the treatment-response curves for intravenous fluid with respect to in-hospital survival and clinical improvement, respectively; panels (g-i) and (j-l) for norepinephrine equivalent dose. For each curve, the other treatment is considered fixed at the model-recommended optimal dose. Darker colors represent stronger magnitude of the effect, blue hues indicate beneficial effects of treatment as opposed to red. The dashed line represent median prediction, dark lines represent 95% CI. Each panel show an example patient. Alt text: Twelve line plots of the change in predicted outcome probability, in percentage points, against simulated dose. The top rows show intravenous fluid and the bottom rows norepinephrine equivalent dose, for in-hospital survival and clinical improvement in turn. Each panel is one example patient, with a dashed median curve and dark lines bounding the 95 percent confidence interval. Blue shading marks beneficial effects and red harmful ones.

### Robustness to design choices

In external validation, severity stratification shows significant variance across cohorts (Figure 6). For clinical improvement, larger recommendation deviations significantly associate with poorer outcomes across strata, peaking for vasopressors in low-severity patients (OR: 1.53, 95% CI: [1.43, 1.63]). For in-hospital survival, fluid deviations in critical patients (OR: 0.93, 95% CI: [0.87, 1.00]) and vasopressor deviations in low-severity patients (OR: 1.08, 95% CI: [0.96, 1.19]) lose statistical significance.

**Figure 6:**
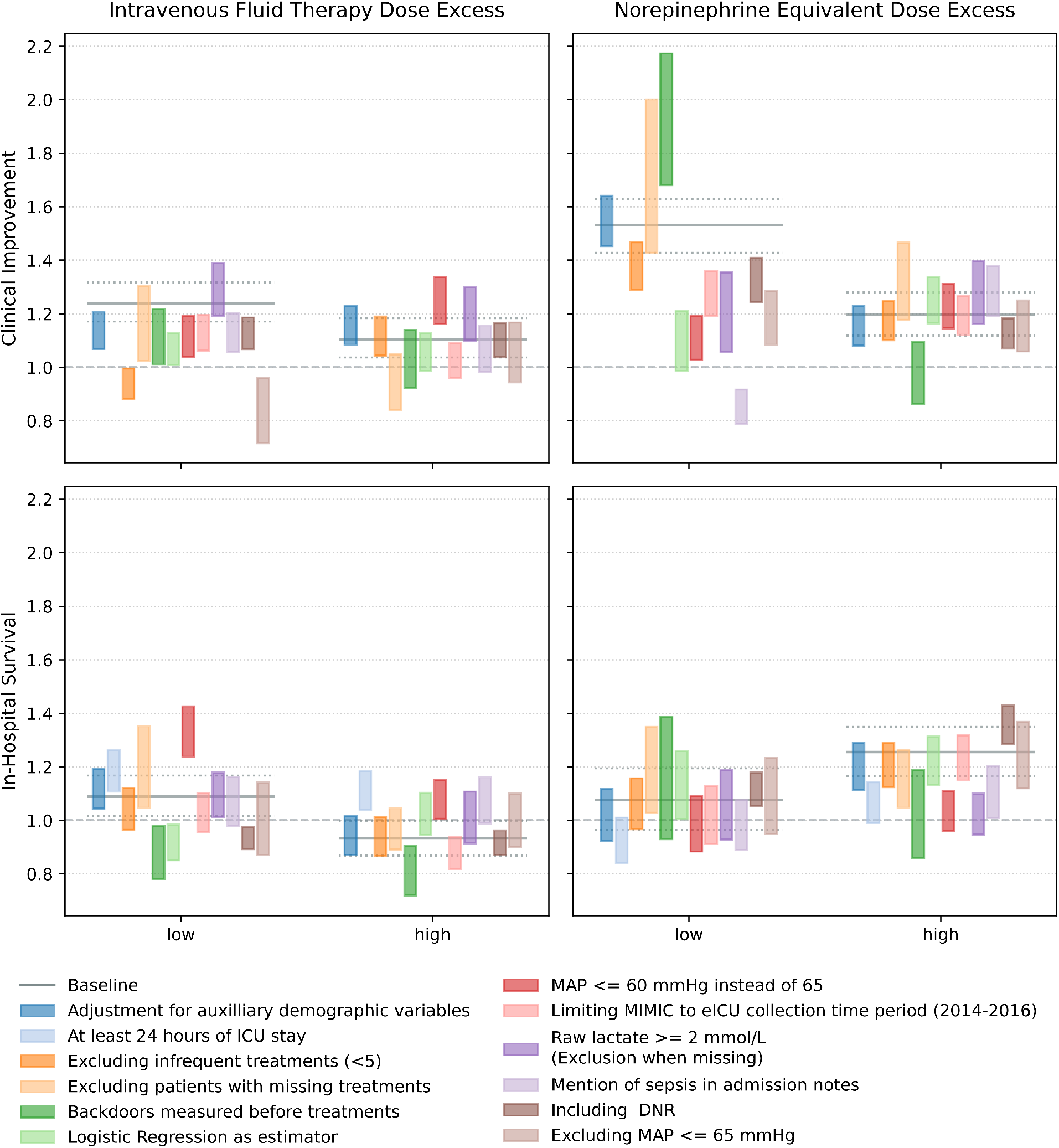
Odds of missing in-hospital survival and clinical improvement with respect to deviation between the observed delivered treatment and the optimal causal suggestion, stratified by severity for different sensitivity analysis. The Y-axis represents odds of missing the target outcome. X-axis represent the categories of severity investigated according to SOFA. Alt text: Grouped plot of the odds of missing each outcome as the delivered dose deviates from the causal recommendation, plotted against SOFA severity category on the horizontal axis and repeated across the sensitivity analysis configurations.

The association between vasopressor deviation and failed clinical improvement was the most stable finding: the odds ratio exceeded 1 in all twelve configurations in which the endpoint could be computed, and the 95% CI excluded 1 in eleven (Table S8). The association with in-hospital survival was less stable, being null in five of the twelve analyses (mention of sepsis 1.03; MAP *≤* 60 mmHg 1.01; raw lactate 1.03; *≥* 24 h ICU stay 0.97; pre-treatment backdoors 1.07). Under pre-treatment backdoors (n = 378) fluid deviation for survival inverted to 0.81 (95% CI: [0.74, 0.87]), while vasopressor deviation for improvement remained robust (OR 1.39). In external validation, requiring an explicit admission sepsis note (n = 969) reduced the causal survival AUROC to 0.64 (95% CI: [0.59, 0.70]); discrimination and effect heterogeneity are nonetheless distinct, since responsiveness rests on the treatment-covariate interaction surface rather than on baseline risk ranking. Restricting external validation to admissions with complete treatment documentation (n = 320) preserved both associations (OR 1.17, 95% CI: [1.08, 1.27] for survival; 1.52, 95% CI: [1.37, 1.64] for improvement). Directional inversions for vasopressors arose only after stratification by severity (Table S10) in sepsis notes in low-severity patients (OR 0.86, 95% CI: [0.79, 0.92]).

The causal model underperformed predictive baselines in external validation in only three configurations, with AUROC gaps between *−*0.03 and *−*0.05 (DNR inclusion for survival; logistic regression; truncated MIMIC training windows for improvement).

Detailed sensitivity specifications and tables are in Tables S8, S9, S10, S7, and S6 (B.3).

## Discussion

Our proposed framework, evaluated on 3,156 ICU admissions across the MIMIC and eICU databases, provides a foundational example of how to introduce this structured causal methodology in intensive care. The causal graph ensures alignment between modeling assumptions and clinical reasoning, while enabling the estimation of hypothetical treatment effects directly from observational data. A notable finding was that the optimal policy space over fluid and vasopressor doses was non-concave, rather than showing the simple U-shape that would point to a single optimal policy. Accordingly, our causal model identifies a set of feasible policies where the clinician retains the final decision-making authority, a property we argue is not only inevitable given the complexity of the problem at hand, but also desirable in clinical practice.

Notably, by explicitly integrating causal reasoning into the modelling framework, the causal model uses approximately 65% fewer variables than conventional predictive approaches. This reduces data requirements and computational complexity, easing deployment across diverse clinical environments while preserving clinically relevant decision support. Together with its comparable predictive performance and robustness under changing conditions, these findings highlight the potential of causal machine learning to support more reliable and generalizable clinical decision-making. These desirable properties are rooted in the process of constructing the causal graph itself. Constructing such a graph requires sustained collaboration between clinicians and data scientists, and so builds the joint teams any clinical AI application depends on. Although the graph is necessarily a simplification and may be wrong, its explicitness lets other researchers scrutinize and improve it: unlike a black-box model, it offers a shared representational language supporting incremental refinement and trust.

This study has several limitations, the first concerning the validation strategy itself. Clinicians escalate vasopressors in patients who are deteriorating, so larger deviations from a model recommending less will tend to mark sicker patients; severity stratification attenuates but cannot remove this, and the deviation analysis should therefore be read as face validity rather than as evidence that following the policy improves outcomes. Interventional validation also remains vulnerable to survivor bias, immortal time bias, and time-zero bias. Second, the causal graph isolates hemodynamic physiology, ignoring concurrent interventions like mechanical ventilation or secondary diagnoses. Third, variable imputation and aggregation may reflect operational biases rather than physiology. Conditioning endpoints on 30-hour survival restricts our estimand to 30-hour survivors, excluding severe early deaths where hemodynamic management is critical. Cohort selection also diverges: eICU excluded transferred patients, unlike MIMIC. Additionally, static modeling oversimplifies time-varying dynamics. We also identified potential fairness concerns, including reduced performance in Hispanic populations and a clinical improvement AUROC gap for female patients. Finally, in the development cohort the model outperformed a constant predictor set at the observed prevalence (Brier 0.17 vs 0.19 for survival), but in external validation it did not (0.12 vs 0.100 for survival; 0.15 vs 0.143 for improvement), indicating miscalibration after transfer. Within-patient comparisons underlying the argmax recommendation are less sensitive to a shift in baseline risk, but the absolute magnitudes in Figure 5 should not be read quantitatively in eICU.

## Conclusions

Constructing a causal graph through multidisciplinary consensus ensures that recommendations are grounded in physiological principles rather than incidental statistical associations. This structural approach enables the *Causal Artificial Intelligence Clinician* to simulate interventional scenarios, providing transparent and clinically interpretable insights for early hemodynamic management. By explicitly encoding domain knowledge, the framework establishes a robust methodology for estimating treatment effects and facilitating evidence-based decision-making in intensive care settings. The dose recommendations themselves remain hypothesis-generating and require prospective evaluation.

**Table 1:**
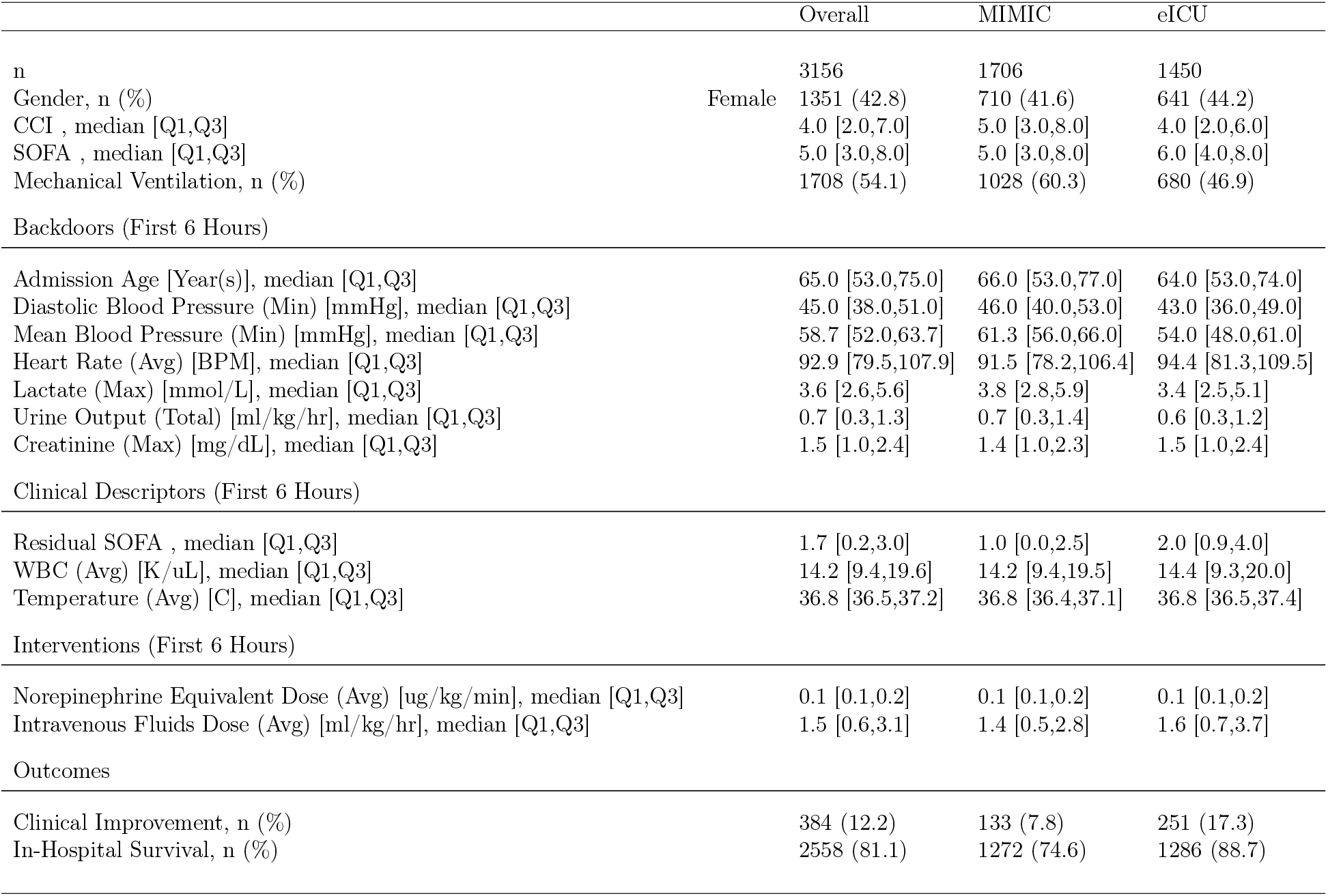
Cohort Description. CCI: Charlson Comorbidity Index, SOFA Score: Sequential Organ Failure Assessment Score, Residual SOFA Score: SOFA score excluding renal and cardiovascular sections. Values refer to observed data before imputation.

## Data Availability

Data used in this research was collected from publicly available databases, the
MIMIC-IV and eICU databases. The procedure to access the MIMIC-IV is
available at: https://physionet.org/content/mimiciv/3.1/. The procedure to access
the eICU is available at: https://physionet.org/content/eicu-crd/2.0/. To recre-
ate the specific datasets used in this study you can use our ETL available at:
https://github.com/IDSIA/causal-ai-clinician

https://physionet.org/content/eicu-crd/2.0/

## Author contributions

G.A.: data curation, formal analysis, investigation, methodology, resources, software, visualization, writing – original draft. L.A.: conceptualization, investigation, methodology, supervision, visualization, writing – review and editing. M.C.: conceptualization, investigation, methodology, supervision, visualization, writing – review and editing. M.Z.: conceptualization, investigation, methodology, supervision, visualization, writing – review and editing. M.C. and M.Z. jointly supervised this work. All authors read and approved the submitted manuscript.

## Acknowledgments

**Artificial intelligence tool use**

Large language models (Google Gemini and Anthropic Claude) were used for language editing, grammar and spelling correction, table and LATEX formatting, and proofreading, and as a technical reference during code development in a manner comparable to conventional programming documentation. Conceptualization, derivation of the causal graph, study design, statistical analysis, interpretation of the results and all conclusions were developed entirely by the authors. No model-generated text was included without author review and revision, and no model is listed as an author. The authors take full responsibility for the integrity, accuracy and originality of the work.

## Data availability

This research used the publicly available MIMIC-IV and eICU Collaborative Research Databases. Access procedures are documented at https://physionet.org/content/mimiciv/3.1/ and https://physionet.org/content/eicu-crd/2.0/. Instructions to rebuild the datasets and reproduce the analysis are available at https://github.com/IDSIA/causal-ai-clinician.

## Ethical approval

The MIMIC-IV dataset is de-identified, and its use for research was approved by the institutional review boards of the Massachusetts Institute of Technology (0403000206) and the Beth Israel Deaconess Medical Center (2001-P-001699/14). Use of the eICU database is exempt from institutional review board approval owing to the retrospective design, the absence of direct patient intervention, and a security schema whose re-identification risk was certified as meeting safe harbor standards by an independent privacy expert (Privacert, Cambridge, MA; Health Insurance Portability and Accountability Act Certification No. 1031219-2). The study was conducted in accordance with the Declaration of Helsinki.

