## Supplementary Material for "The Causal Artificial Intelligence Clinician for early hemodynamic management of septic shock in ICU"

---

#### A. List of Abbreviations

- **AI**: Artificial Intelligence
- **ATE**: Average Treatment Effect
- **AUPRC**: Area Under the Precision-Recall Curve
- **AUROC**: Area Under the Receiver Operating Characteristic curve
- **CATE**: Conditional Average Treatment Effect
- **CI**: Confidence Interval
- **CCI**: Charlson Comorbidity Index
- **DAG**: Directed Acyclic Graph
- **DNR**: Do Not Resuscitate
- **ED**: Emergency Department
- **eICU**: eICU Collaborative Research Database
- **EHR**: Electronic Health Records

*Corresponding author
[1]These authors contributed equally to this work.

- **ER**: Emergency Room
- **ETT**: Effect of the Treatment on the Treated
- **GCS**: Glasgow Coma Scale
- **ICU**: Intensive Care Unit
- **IQR**: Interquartile Range
- **LMIC**: Low- and Middle-Income Countries
- **MAR**: Missing At Random
- **MIMIC**: Medical Information Mart for Intensive Care
- **ML**: Machine Learning
- **RCT**: Randomized Controlled Trial
- **SCM**: Structural Causal Model
- **SOFA**: Sequential Organ Failure Assessment
- **STEM**: Science, Technology, Engineering, and Mathematics
- **WBC**: White Blood Cell

#### B. Cohort details and commentary

Following the application of inclusion and exclusion criteria (Fig. S2), we obtained a final cohort comprising 3,156 intensive care unit admissions, split in 1,706 from the MIMIC and 1,450 from the eICU. Overall, our cohort exhibited a male majority (57.2 percent) and a high rate of invasive mechanical ventilation (54.1 percent). Baseline indicators demonstrated substantial comorbidity, with a median Charlson Comorbidity Index (CCI) of 4 (interquartile range [IQR] 2-7), and elevated admission severity, marked by a median SOFA score of 5 (IQR 3-8). Outcome assessment revealed an overall in-hospital survival rate of 81.1 percent with 12.2 percent of patients achieving clinical improvement (2-points reduction in SOFA score). Significant clinical heterogeneity was present between the training and external validation dataset. Patients sourced from the MIMIC database generally exhibited more severe conditions (SOFA, odds ratio [OR] 1.38, 95% CI: [1.23, 1.57]) despite a lower unadjusted median SOFA score (MIMIC median 5, IQR 3-8 vs. eICU median 6, IQR 4-8). Furthermore, the MIMIC sub-cohort presented with higher comorbidity (CCI, OR 1.19, 95% CI: [1.11, 1.32]) and higher vasopressor requirements (OR 1.19, 95% CI: [1.10, 1.38]). MIMIC patients had a higher likelihood of poor outcomes, including lower in-hospital survival (74.6 vs. 88.7 percent; OR 0.79, 95% CI 0.65-0.87) and lower clinical improvement rates (7.8 vs. 17.3 percent; OR 0.88, 95% CI: [0.79, 0.93]). Conversely, patients in the eICU sub-cohort recorded higher non-cardiac and non-renal SOFA components (residual SOFA, OR 0.71, 95% CI: [0.58, 0.80]) and higher volumes of intravenous fluid intake (OR 0.92, 95% CI: [0.89, 0.95]). Comprehensive descriptors are detailed in Table S1 and Table S2.

| | | Overall | MIMIC | eICU |
|---|---|---|---|---|
| n | | 3156 | 1706 | 1450 |
| **Admission Age** [Year(s)], median [Q1,Q3] | | 65.0 [53.0,75.0] | 66.0 [53.0,77.0] | 64.0 [53.0,74.0] |
| Weight [Kgs], median [Q1,Q3] | | 79.0 [66.0,95.7] | 78.0 [65.8,93.9] | 80.4 [66.2,98.9] |
| CCI , median [Q1,Q3] | | 4.0 [2.0,7.0] | 5.0 [3.0,8.0] | 4.0 [2.0,6.0] |
| Gender, n (%) | Female | 1351 (42.8) | 710 (41.6) | 641 (44.2) |
| Hypertension, n (%) | | 1267 (40.1) | 562 (32.9) | 705 (48.6) |
| Infection Location | | | | |
| Abdominal Infection, n (%) | | 123 (3.9) | 95 (5.6) | 28 (1.9) |
| Respiratory Infection, n (%) | | 851 (27.0) | 481 (28.2) | 370 (25.5) |
| CNS Infection, n (%) | | 21 (0.7) | 15 (0.9) | 6 (0.4) |
| Urinary Infection, n (%) | | 421 (13.3) | 203 (11.9) | 218 (15.0) |
| Scores (First 6 Hours) | | | | |
| SOFA , median [Q1,Q3] | | 5.0 [3.0,8.0] | 5.0 [3.0,8.0] | 6.0 [4.0,8.0] |
| **Residual SOFA** , median [Q1,Q3] | | 1.7 [0.2,3.0] | 1.0 [0.0,2.5] | 2.0 [0.9,4.0] |
| Vitals (First 6 Hours) | | | | |
| GCS (Min), median [Q1,Q3] | | 15.0 [13.9,15.0] | 15.0 [15.0,15.0] | 14.0 [9.5,15.0] |
| Respiratory Rate (Avg) [BrPM], median [Q1,Q3] | | 20.5 [17.7,23.9] | 20.5 [17.8,23.7] | 20.5 [17.5,24.1] |
| **Heart Rate** (Avg) [BPM], median [Q1,Q3] | | 92.9 [79.5,107.9] | 91.5 [78.2,106.4] | 94.4 [81.3,109.5] |
| **Mean Blood Pressure** (Min) [mmHg], median [Q1,Q3] | | 58.7 [52.0,63.7] | 61.3 [56.0,66.0] | 54.0 [48.0,61.0] |
| **Diastolic Blood Pressure** (Min) [mmHg], median [Q1,Q3] | | 45.0 [38.0,51.0] | 46.0 [40.0,53.0] | 43.0 [36.0,49.0] |
| **Urine Output** (Total) [ml/kg/hr], median [Q1,Q3] | | 0.7 [0.3,1.3] | 0.7 [0.3,1.4] | 0.6 [0.3,1.2] |
| **Temperature** (Avg) [C], median [Q1,Q3] | | 36.8 [36.5,37.2] | 36.8 [36.4,37.1] | 36.8 [36.5,37.4] |
| SpO2 (Min) [%], median [Q1,Q3] | | 94.0 [91.0,97.0] | 95.0 [92.0,98.0] | 93.0 [90.0,96.0] |
| Laboratory (First 6 Hours) | | | | |
| Bilirubin (Max) [mg/dL], median [Q1,Q3] | | 0.7 [0.4,1.4] | 0.7 [0.4,1.6] | 0.7 [0.4,1.3] |
| Hemoglobin (Min) [g/dL], median [Q1,Q3] | | 10.6 [8.7,12.3] | 10.5 [8.7,12.3] | 10.7 [9.0,12.4] |
| Platelets (Min), median [Q1,Q3] | | 180.0 [120.0,255.0] | 176.5 [118.0,256.0] | 186.0 [125.0,253.5] |
| **WBC** (Avg) [K/uL], median [Q1,Q3] | | 14.2 [9.4,19.6] | 14.2 [9.4,19.5] | 14.4 [9.3,20.0] |
| **Creatinine** (Max) [mg/dL], median [Q1,Q3] | | 1.5 [1.0,2.4] | 1.4 [1.0,2.3] | 1.5 [1.0,2.4] |
| **Lactate** (Max) [mmol/L], median [Q1,Q3] | | 3.6 [2.6,5.6] | 3.8 [2.8,5.9] | 3.4 [2.5,5.1] |
| Interventions | | | | |
| Norepinephrine Equivalent Dose (Avg) [ug/kg/min], median [Q1,Q3] | | 0.1 [0.1,0.2] | 0.1 [0.1,0.2] | 0.1 [0.1,0.2] |
| Intravenous Fluids Dose (Avg) [ml/kg/hr], median [Q1,Q3] | | 1.5 [0.6,3.1] | 1.4 [0.5,2.8] | 1.6 [0.7,3.7] |
| Mechanical Ventilation, n (%) | | 1708 (54.1) | 1028 (60.3) | 680 (46.9) |
| Others | | | | |
| Admission Time, n (%) | 00-06 | 765 (24.2) | 341 (20.0) | 424 (29.2) |
| | 06-12 | 581 (18.4) | 314 (18.4) | 267 (18.4) |
| | 12-18 | 783 (24.8) | 503 (29.5) | 280 (19.3) |
| | 18-24 | 1027 (32.5) | 548 (32.1) | 479 (33.0) |
| Admission Year, n (%) | 2011 - 2014 | 400 (12.7) | 400 (23.4) | 0 (0.0) |
| | 2014 - 2017 | 2119 (67.1) | 669 (39.2) | 1450 (100.0) |
| | 2017 - 2020 | 595 (18.9) | 595 (34.9) | 0 (0.0) |
| | 2020 - 2023 | 42 (1.3) | 42 (2.5) | 0 (0.0) |
| Unit, n (%) | Cardiac | 550 (17.4) | 245 (14.4) | 305 (21.0) |
| | Medical-Surgical | 2267 (71.8) | 1197 (70.2) | 1070 (73.8) |
| | Neuro-Trauma | 339 (10.7) | 264 (15.5) | 75 (5.2) |
| Post-vasotherapy MBP readings , n (%) | | 397 (12.6) | 370 (21.7) | 27 (1.9) |
| Outcomes | | | | |
| Clinical Improvement, n (%) | | 384 (12.2) | 133 (7.8) | 251 (17.3) |
| In-Hospital Survival, n (%) | | 2558 (81.1) | 1272 (74.6) | 1286 (88.7) |

Table S1: Full set of features accessible to the predictive model plus outcomes. CCI: Charlson Comorbidity Index, SOFA Score: Sequential Organ Failure Assessment Score, Residual SOFA Score: SOFA score excluding renal and cardiovascular sections,GCS: Glasgow Coma Scale, WBC: White Blood Cell Count, SpO2: Blood oxygen saturation, MBP: Mean Blood Pressure. In bold variables were used by the causal model.

| **Feature** | **Odds** median [95CI] |
|---|---|
| Admission Year:2017 - 2020 | 1.67 [1.33,2.63] |
| Admission Year:2011 - 2014 | 1.49 [1.25,2.14] |
| Mechanical Ventilation | 1.46 [1.23,2.03] |
| GCS | 1.47 [1.27,1.64] |
| SOFA | 1.38 [1.23,1.57] |
| Post-Vasotherapy MBP | 1.30 [1.16,1.63] |
| Clinical Stability | 1.28 [1.15,1.60] |
| Unit:Neuro-Trauma | 1.23 [1.12,1.49] |
| Hemoglobin | 1.23 [1.08,1.30] |
| Norepinephrine Equivalent Dose | 1.19 [1.10,1.38] |
| CCI | 1.19 [1.11,1.32] |
| Urine Output | 1.16 [1.07,1.33] |
| Mean Blood Pressure | 1.17 [1.10,1.23] |
| Admission Time:12-18 | 1.14 [1.08,1.29] |
| Abdominal Infection | 1.07 [1.04,1.14] |
| Lactate | 1.07 [0.97,1.15] |
| Respiratory Infection | 1.05 [1.03,1.10] |
| Gender | 1.04 [1.02,1.09] |
| Admission Year:2020 - 2023 | 1.04 [1.02,1.08] |
| SpO2 | 1.04 [1.02,1.07] |
| Admission Time:06-12 | 1.02 [1.00,1.06] |
| CNS Infection | 1.01 [1.00,1.02] |
| Heart Rate | 1.00 [0.99,1.01] |
| Platelets | 1.00 [1.00,1.00] |
| Weight | 0.99 [0.99,1.00] |
| WBC | 0.99 [0.96,1.02] |
| Respiratory Rate | 0.99 [0.95,1.07] |
| Bilirubin | 0.97 [0.94,1.13] |
| Admission Age | 0.99 [0.97,1.00] |
| Admission Time:18-24 | 0.98 [0.95,1.00] |
| Urinary Infection | 0.96 [0.92,0.98] |
| Intravenous Fluids Dose | 0.92 [0.89,0.95] |
| Unit:Medical-Surgical | 0.91 [0.85,0.95] |
| Diastolic Blood Pressure | 0.90 [0.85,0.96] |
| Clinical Improvement | 0.88 [0.79,0.93] |
| Unit:Cardiac | 0.88 [0.77,0.94] |
| Admission Time:00-06 | 0.87 [0.76,0.92] |
| Creatinine | 0.78 [0.69,0.88] |
| In-Hospital Survival | 0.79 [0.65,0.87] |
| Hypertension | 0.78 [0.62,0.87] |
| Residual SOFA | 0.71 [0.58,0.80] |
| Temperature | 0.67 [0.61,0.76] |
| Admission Year:2014 - 2017 | 0.38 [0.17,0.58] |

Table S2: Odds of a patient belonging to the MIMIC cohort as against eICU. Odds ratios are multivariable, mutually adjusted for every covariate listed in the table.

| **Feature** | **MIMIC (%)** | **eICU (%)** |
|---|---|---|
| Admission Age | 0.0 | 0.0 |
| Weight | 0.0 | 0.0 |
| CCI | 0.0 | 0.0 |
| Gender | 0.0 | 0.0 |
| Hypertension | 0.0 | 0.0 |
| SOFA | 0.4 | 0.0 |
| Residual SOFA | 0.4 | 0.0 |
| GCS | 1.8 | 20.3 |
| Respiratory Rate | 0.1 | 0.9 |
| Heart Rate | 0.2 | 0.1 |
| Mean Blood Pressure | 0.2 | 0.0 |
| Diastolic Blood Pressure | 0.2 | 0.5 |
| Urine Output | 10.6 | 31.9 |
| Temperature | 5.6 | 1.9 |
| SpO2 | 0.4 | 2.0 |
| Abdominal Infection | 0.0 | 0.0 |
| Respiratory Infection | 0.0 | 0.0 |
| CNS Infection | 0.0 | 0.0 |
| Urinary Infection | 0.0 | 0.0 |
| Bilirubin | 21.8 | 52.7 |
| Hemoglobin | 0.5 | 34.8 |
| Platelets | 0.5 | 37.7 |
| WBC | 0.5 | 37.7 |
| Creatinine | 0.4 | 30.8 |
| Lactate | 28.3 | 43.4 |
| Norepinephrine Equivalent Dose | 29.6 | 77.5 |
| Intravenous Fluids Dose | 0.0 | 0.0 |
| Mechanical Ventilation | 0.0 | 0.0 |
| Admission Time | 0.0 | 0.0 |
| Admission Year | 0.0 | 0.0 |
| Unit | 0.0 | 0.0 |
| Post-Vasotherapy MBP | 0.0 | 0.0 |
| Clinical Stability | 0.0 | 0.0 |
| Clinical Improvement | 0.0 | 0.0 |
| In-Hospital Survival | 0.0 | 0.0 |

Table S3: Missing rates across MIMIC and eICU variables.

##### B.1. Fairness Analysis Details

| Category | Subgroup | Causal | Predictive |
|---|---|---|---|
| Gender | Female | -0.00 [-0.09,0.08] | 0.06 [-0.03,0.14] |
| Race and Ethnicity | Asian | | |
| | Black | 0.00 [-0.13,0.12] | 0.01 [-0.12,0.12] |
| | Hispanic | -0.17 [-0.36,0.01] | -0.15 [-0.33,0.01] |
| | Other | -0.09 [-0.28,0.07] | -0.07 [-0.24,0.08] |
| | White | -0.03 [-0.14,0.08] | -0.04 [-0.15,0.06] |

(a)

| Category | Subgroup | Causal | Predictive |
|---|---|---|---|
| Gender | Female | -0.07 [-0.14,-0.00] | -0.04 [-0.11,0.03] |
| Race and Ethnicity | Asian | -0.17 [-0.78,0.28] | -0.13 [-0.79,0.27] |
| | Black | -0.08 [-0.33,0.06] | -0.11 [-0.36,0.04] |
| | Hispanic | -0.09 [-0.38,0.11] | -0.03 [-0.32,0.14] |
| | Other | -0.06 [-0.34,0.10] | -0.10 [-0.36,0.08] |
| | White | -0.08 [-0.32,0.03] | -0.13 [-0.35,-0.01] |

(b)

Table S4: AUROC parity test for (S4a) In-Hospital Survival and (S4b) Clinical Improvement. Values presented as Median [95% CI] based on simultaneous Hsu's confidence intervals method. An interval whose upper bound is exactly 0.000 is not significant at the 5% level.

##### B.2. Cross-Validation and External Validation Details

| | Clinical Improvement | | In-Hospital Survival | |
|---|---|---|---|---|
| | MIMIC | eICU | MIMIC | eICU |
| Causal | 0.77 [0.67,0.81] | 0.70 [0.66,0.73] | 0.73 [0.71,0.75] | 0.72 [0.67,0.76] |
| Predictive | 0.77 [0.73,0.85] | 0.68 [0.65,0.72] | 0.77 [0.74,0.79] | 0.74 [0.70,0.78] |

(a)

| | Clinical Improvement | | In-Hospital Survival | |
|---|---|---|---|---|
| | MIMIC | eICU | MIMIC | eICU |
| Causal | 0.25 [0.14,0.31] | 0.32 [0.26,0.37] | 0.89 [0.87,0.89] | 0.95 [0.93,0.96] |
| Predictive | 0.22 [0.20,0.50] | 0.32 [0.27,0.37] | 0.90 [0.88,0.91] | 0.95 [0.94,0.96] |

(b)

| | Clinical Improvement | | In-Hospital Survival | |
|---|---|---|---|---|
| | MIMIC | eICU | MIMIC | eICU |
| Causal | 0.07 [0.07,0.07] | 0.15 [0.13,0.16] | 0.17 [0.16,0.17] | 0.12 [0.11,0.12] |
| Predictive | 0.07 [0.06,0.07] | 0.15 [0.13,0.16] | 0.16 [0.15,0.16] | 0.11 [0.11,0.12] |

(c)

Table S5: Predictive performance.(S5a) AUROC, (S5b) AUPRC and (S5c) Brier Score in cross-validation (MIMIC) and external validation (eICU) for the studys' endpoints. Values presented as Median [95% CI].

##### *B.3. Sensitivity Analysis Details and Commentary*

| **Criteria** | | n | **In-Hospital Survival** % | **Causal** median [95CI] | **Predictive** median [95CI] | **Gap** median [95CI] |
|---|---|---|---|---|---|---|
| Baseline | MIMIC | 1706 | .75 | **0.73 [0.71,0.75]** | **0.77 [0.74,0.79]** | **-0.04 [-0.05,-0.01]** |
| | eICU | 1450 | .89 | 0.72 [0.67,0.76] | 0.74 [0.70,0.78] | -0.02 [-0.04,-0.00] |
| Including DNR | MIMIC | 1797 | .74 | 0.72 [0.70,0.77] | 0.75 [0.74,0.78] | -0.05 [-0.06,0.04] |
| | eICU | 2076 | .75 | **0.73 [0.71,0.76]** | **0.76 [0.74,0.79]** | **-0.03 [-0.04,-0.02]** |
| Excluding MAP $\leq$ 65 mmHg | MIMIC | 1197 | .71 | 0.71 [0.70,0.72] | 0.75 [0.71,0.77] | -0.03 [-0.08,-0.00] |
| | eICU | 320 | .81 | 0.72 [0.64,0.79] | 0.75 [0.68,0.81] | -0.03 [-0.07,0.00] |
| Mention of sepsis in admission notes | MIMIC | 271 | .79 | **0.50 [0.42,0.61]** | **0.59 [0.50,0.68]** | **-0.09 [-0.11,-0.02]** |
| | eICU | 969 | .89 | 0.64 [0.59,0.70] | 0.67 [0.61,0.72] | -0.02 [-0.07,0.03] |
| MAP $\leq$ 60 mmHg instead of 65 | MIMIC | 1466 | .73 | 0.74 [0.67,0.77] | 0.76 [0.65,0.79] | -0.02 [-0.10,0.12] |
| | eICU | 1179 | .88 | 0.75 [0.71,0.79] | 0.76 [0.72,0.81] | -0.01 [-0.04,0.01] |
| Raw lactate $\geq$ 2 mmol/L | MIMIC | 1221 | .71 | 0.73 [0.71,0.74] | 0.75 [0.73,0.76] | -0.02 [-0.03,-0.00] |
| | eICU | 811 | .87 | 0.74 [0.69,0.78] | 0.74 [0.70,0.79] | -0.01 [-0.03,0.02] |
| Adjustment for auxilliary demographic variables | MIMIC | 1706 | .75 | 0.75 [0.66,0.80] | 0.76 [0.72,0.79] | -0.03 [-0.14,0.04] |
| | eICU | 1450 | .89 | 0.72 [0.67,0.76] | 0.74 [0.70,0.78] | -0.02 [-0.04,-0.00] |
| At least 24 hours of ICU stay | MIMIC | 1812 | .74 | 0.74 [0.72,0.78] | 0.79 [0.73,0.81] | -0.02 [-0.08,0.03] |
| | eICU | 1547 | .88 | 0.72 [0.68,0.76] | 0.74 [0.70,0.78] | -0.03 [-0.05,-0.00] |
| Logistic Regression as estimator | MIMIC | 1706 | .75 | 0.72 [0.65,0.74] | 0.73 [0.70,0.81] | -0.02 [-0.10,0.02] |
| | eICU | 1450 | .89 | 0.70 [0.65,0.74] | 0.72 [0.68,0.77] | -0.03 [-0.05,0.00] |
| Excluding infrequent treatments (<5) | MIMIC | 1706 | .75 | 0.72 [0.70,0.78] | 0.77 [0.74,0.79] | -0.05 [-0.09,0.01] |
| | eICU | 1450 | .89 | 0.72 [0.67,0.76] | 0.74 [0.70,0.78] | -0.02 [-0.04,-0.00] |
| Excluding patients with missing treatments | MIMIC | 926 | .67 | 0.71 [0.65,0.75] | 0.69 [0.68,0.77] | -0.00 [-0.12,0.06] |
| | eICU | 320 | .81 | 0.71 [0.64,0.78] | 0.74 [0.67,0.80] | -0.03 [-0.06,0.01] |
| Backdoors measured before treatments | MIMIC | 1015 | .7 | 0.73 [0.70,0.74] | 0.73 [0.72,0.78] | -0.03 [-0.05,0.01] |
| | eICU | 378 | .87 | 0.70 [0.61,0.78] | 0.73 [0.65,0.81] | -0.03 [-0.06,-0.00] |
| Limiting MIMIC to eICU collection time period (2014-2016) | MIMIC | 550 | .75 | 0.74 [0.72,0.78] | 0.76 [0.73,0.78] | -0.02 [-0.06,0.04] |
| | eICU | 1450 | .89 | 0.72 [0.67,0.76] | 0.73 [0.69,0.77] | -0.02 [-0.04,0.00] |

Table S6: Sensitivity Analysis for the in-hospital survival endpoint: AUROC for different exclusion and inclusion criteria and choices of backdoors. The gap column reports the median of the paired bootstrap differences between the causal and the predictive model AUROC; it is therefore not the difference of the two reported medians and may carry the opposite sign.

| **Criteria** | | n | **Clinical Improvement** % | **Causal** median [95CI] | **Predictive** median [95CI] | **Gap** median [95CI] |
|---|---|---|---|---|---|---|
| Baseline | MIMIC | 1706 | .08 | 0.77 [0.67,0.81] | 0.77 [0.73,0.85] | -0.09 [-0.10,0.09] |
| | eICU | 1450 | .17 | 0.70 [0.66,0.73] | 0.68 [0.65,0.72] | 0.01 [-0.02,0.04] |
| Including DNR | MIMIC | 1797 | .08 | 0.74 [0.69,0.81] | 0.80 [0.78,0.81] | -0.06 [-0.10,0.00] |
| | eICU | 2076 | .16 | 0.69 [0.66,0.72] | 0.69 [0.66,0.71] | 0.00 [-0.02,0.02] |
| Excluding MAP $\leq$ 65 mmHg | MIMIC | 1197 | .09 | 0.75 [0.61,0.82] | 0.76 [0.70,0.81] | -0.01 [-0.18,0.05] |
| | eICU | 320 | .24 | 0.72 [0.65,0.78] | 0.64 [0.57,0.71] | 0.07 [0.00,0.14] |
| Mention of sepsis in admission notes | MIMIC | 271 | .09 | 0.75 [0.70,0.88] | 0.76 [0.68,0.94] | -0.04 [-0.18,0.08] |
| | eICU | 969 | .18 | 0.66 [0.61,0.70] | 0.66 [0.62,0.71] | -0.01 [-0.03,0.02] |
| MAP $\leq$ 60 mmHg instead of 65 | MIMIC | 1466 | .09 | 0.75 [0.69,0.78] | 0.78 [0.72,0.86] | -0.05 [-0.09,0.04] |
| | eICU | 1179 | .17 | 0.70 [0.66,0.73] | 0.68 [0.65,0.72] | 0.01 [-0.02,0.04] |
| Raw lactate $\geq$ 2 mmol/L | MIMIC | 1221 | .08 | 0.70 [0.65,0.78] | 0.76 [0.75,0.83] | -0.10 [-0.15,0.04] |
| | eICU | 811 | .18 | 0.68 [0.63,0.72] | 0.70 [0.66,0.75] | -0.03 [-0.05,0.00] |
| Adjustment for auxilliary demographic variables | MIMIC | 1706 | .08 | 0.75 [0.70,0.80] | 0.84 [0.72,0.87] | -0.07 [-0.14,0.09] |
| | eICU | 1450 | .17 | 0.70 [0.66,0.73] | 0.68 [0.65,0.72] | 0.01 [-0.01,0.04] |
| Logistic Regression as estimator | MIMIC | 1706 | .08 | **0.70 [0.68,0.81]** | **0.80 [0.74,0.82]** | **-0.04 [-0.11,-0.01]** |
| | eICU | 1450 | .17 | **0.68 [0.64,0.71]** | **0.73 [0.70,0.76]** | **-0.05 [-0.08,-0.03]** |
| Excluding infrequent treatments (<5) | MIMIC | 1706 | .08 | 0.72 [0.71,0.75] | 0.77 [0.71,0.84] | -0.02 [-0.13,0.01] |
| | eICU | 1450 | .17 | 0.70 [0.66,0.73] | 0.68 [0.65,0.72] | 0.01 [-0.02,0.04] |
| Excluding patients with missing treatments | MIMIC | 926 | .09 | 0.73 [0.69,0.82] | 0.77 [0.72,0.88] | -0.03 [-0.17,0.05] |
| | eICU | 320 | .24 | 0.73 [0.67,0.79] | 0.70 [0.63,0.76] | 0.03 [-0.01,0.08] |
| Backdoors measured before treatments | MIMIC | 1015 | .06 | 0.71 [0.66,0.79] | 0.80 [0.67,0.85] | -0.07 [-0.14,0.04] |
| | eICU | 378 | .13 | 0.66 [0.58,0.73] | 0.73 [0.66,0.80] | -0.07 [-0.14,0.00] |
| Limiting MIMIC to eICU collection time period (2014-2016) | MIMIC | 550 | .06 | 0.64 [0.54,0.80] | 0.70 [0.56,0.78] | 0.05 [-0.25,0.10] |
| | eICU | 1450 | .17 | **0.64 [0.61,0.68]** | **0.69 [0.65,0.72]** | **-0.04 [-0.07,-0.02]** |

Table S7: Sensitivity Analysis for the clinical improvement endpoint: AUROC for different exclusion and inclusion criteria and choices of backdoors. The gap column reports the median of the paired bootstrap differences between the causal and the predictive model AUROC; it is therefore not the difference of the two reported medians and may carry the opposite sign. Sensitivity analyses with shorter outcome horizons (see Table S6) were not computed, as insufficient time had elapsed to recompute the SOFA score.

| Criteria | Therapy | **Lack of In-Hospital Survival** median [95CI] | **Lack of Clinical Improvement** median [95CI] |
|---|---|---|---|
| Baseline | Norepinephrine Equivalent | 1.16 [1.10,1.24] | 1.26 [1.21,1.32] |
| | Intravenous Fluid Therapy | 1.02 [0.96,1.07] | 1.12 [1.08,1.18] |
| Including DNR | Norepinephrine Equivalent | 1.16 [1.12,1.21] | 1.20 [1.16,1.24] |
| | Intravenous Fluid Therapy | 0.95 [0.92,0.99] | 1.07 [1.03,1.11] |
| Excluding MAP $\leq$ 65 mmHg | Norepinephrine Equivalent | 1.23 [1.13,1.34] | 1.18 [1.10,1.26] |
| | Intravenous Fluid Therapy | 1.03 [0.94,1.12] | 0.98 [0.90,1.07] |
| Mention of sepsis in admission notes | Norepinephrine Equivalent | 1.03 [0.96,1.10] | 1.05 [1.00,1.10] |
| | Intravenous Fluid Therapy | 1.10 [1.03,1.18] | 1.06 [1.01,1.12] |
| MAP $\leq$ 60 mmHg instead of 65 | Norepinephrine Equivalent | 1.01 [0.95,1.08] | 1.17 [1.11,1.23] |
| | Intravenous Fluid Therapy | 1.19 [1.13,1.25] | 1.14 [1.09,1.20] |
| Raw lactate $\geq$ 2 mmol/L (Exclusion when missing) | Norepinephrine Equivalent | 1.03 [0.97,1.10] | 1.27 [1.18,1.37] |
| | Intravenous Fluid Therapy | 1.06 [0.99,1.12] | 1.18 [1.11,1.25] |
| Adjustment for auxilliary demographic variables | Norepinephrine Equivalent | 1.10 [1.04,1.16] | 1.27 [1.22,1.33] |
| | Intravenous Fluid Therapy | 1.03 [0.98,1.09] | 1.12 [1.07,1.17] |
| At least 24 hours of ICU stay | Norepinephrine Equivalent | 0.97 [0.92,1.03] | |
| | Intravenous Fluid Therapy | 1.15 [1.10,1.21] | |
| Logistic Regression as estimator | Norepinephrine Equivalent | 1.13 [1.07,1.20] | 1.26 [1.19,1.33] |
| | Intravenous Fluid Therapy | 0.97 [0.92,1.02] | 1.07 [1.02,1.11] |
| Excluding infrequent treatments (<5) | Norepinephrine Equivalent | 1.11 [1.06,1.17] | 1.23 [1.18,1.28] |
| | Intravenous Fluid Therapy | 1.00 [0.95,1.06] | 1.00 [0.96,1.05] |
| Excluding patients with missing treatments | Norepinephrine Equivalent | 1.17 [1.08,1.27] | 1.52 [1.37,1.64] |
| | Intravenous Fluid Therapy | 1.04 [0.97,1.13] | 1.03 [0.93,1.11] |
| Backdoors measured before treatments | Norepinephrine Equivalent | 1.07 [0.94,1.26] | 1.39 [1.22,1.54] |
| | Intravenous Fluid Therapy | 0.81 [0.74,0.87] | 1.06 [0.98,1.16] |

| | | | |
|---|---|---|---|
| Limiting MIMIC to eICU collection time period (2014-2016) | Norepinephrine Equivalent | 1.11 [1.05,1.17] | 1.23 [1.17,1.28] |
| | Intravenous Fluid Therapy | 0.98 [0.93,1.04] | 1.06 [1.01,1.11] |

Table S8: Odds of missing each outcome per 1-SD increase in the quantile-normalized deviation between the delivered and the model-recommended dose, in the eICU, for each sensitivity configuration, adjusted for severity (SOFA). Odds are presented as median and 95% CI. A blank cell indicates a configuration in which the endpoint could not be computed.

| Criteria | Severity | Norepinephrine Equivalent Dose Excess | Intravenous Fluid Therapy |
|---|---|---|---|
| Baseline | low | 1.08 [0.96-1.19] | 1.09 [1.02-1.17] |
| | high | 1.25 [1.17-1.35] | 0.93 [0.87-1.00] |
| Including DNR | low | 1.11 [1.05-1.18] | **0.93 [0.89-0.98]** |
| | high | 1.35 [1.28-1.43] | **0.92 [0.87-0.96]** |
| Excluding MAP <= 65 mmHg | low | 1.08 [0.95-1.23] | 0.99 [0.87-1.14] |
| | high | 1.24 [1.12-1.37] | 0.99 [0.90-1.10] |
| Mention of sepsis in admission notes | low | 0.99 [0.89-1.08] | 1.07 [0.98-1.16] |
| | high | 1.10 [1.01-1.20] | 1.07 [0.99-1.16] |
| MAP <= 60 mmHg instead of 65 | low | 0.98 [0.88-1.09] | 1.33 [1.24-1.43] |
| | high | 1.04 [0.96-1.11] | 1.07 [1.01-1.15] |
| Raw lactate >= 2 mmol/L (Exclusion when missing) | low | 1.06 [0.93-1.19] | 1.10 [1.01-1.18] |
| | high | 1.02 [0.95-1.10] | 1.00 [0.91-1.11] |
| Adjustment for auxilliary demographic variables | low | 1.02 [0.92-1.12] | 1.12 [1.04-1.19] |
| | high | 1.20 [1.11-1.29] | 0.94 [0.87-1.02] |
| At least 24 hours of ICU stay | low | 0.93 [0.84-1.01] | 1.18 [1.11-1.26] |
| | high | 1.06 [0.99-1.14] | 1.11 [1.04-1.18] |
| Logistic Regression as estimator | low | 1.13 [1.00-1.26] | **0.92 [0.85-0.98]** |
| | high | 1.22 [1.13-1.31] | 1.01 [0.95-1.10] |
| Excluding infrequent treatments (<5) | low | 1.06 [0.97-1.16] | 1.03 [0.96-1.12] |
| | high | 1.20 [1.12-1.29] | 0.94 [0.87-1.01] |
| Excluding patients with missing treatments | low | 1.15 [1.03-1.35] | 1.17 [1.05-1.35] |
| | high | 1.15 [1.05-1.26] | 0.96 [0.89-1.04] |
| Backdoors measured before treatments | low | 1.16 [0.93-1.39] | **0.89 [0.78-0.98]** |
| | high | 0.96 [0.86-1.19] | **0.82 [0.72-0.90]** |
| Limiting MIMIC to eICU collection time period (2014-2016) | low | 1.02 [0.91-1.13] | 1.03 [0.95-1.10] |
| | high | 1.23 [1.15-1.32] | **0.87 [0.82-0.94]** |

Table S9: Quantile odds between dose excesses and lack of in-hospital mortality in the eICU for different selection criteria after stratifying for severity (SOFA). Odds are presented as median and 95% CI. Odds significantly lower than 1 are reported in bold.

Please note that the clinical improvement endpoint could not be computed in the "At least 24 hours of ICU stay" sensitivity analysis since reducing the ICU length of stay from 30H to 24H does not allow for the estimation of a new 24H SOFA score without using data from the first 6 hours of ICU admission. This affects tables S7, S8 and S10. The same consideration does not apply for the survival endpoint.

| Criteria | Severity | Norepinephrine Equivalent Dose Excess | Intravenous Fluid Therapy |
|---|---|---|---|
| Baseline | low<br>high | 1.53 [1.43-1.63]<br>1.20 [1.12-1.28] | 1.24 [1.17-1.32]<br>1.10 [1.04-1.18] |
| Including DNR | low<br>high | 1.32 [1.24-1.41]<br>1.13 [1.07-1.18] | 1.12 [1.07-1.19]<br>1.11 [1.04-1.17] |
| Excluding MAP <= 65 mmHg | low<br>high | 1.17 [1.08-1.28]<br>1.15 [1.06-1.25] | **0.79 [0.72-0.96]**<br>1.06 [0.94-1.17] |
| Mention of sepsis in admission notes | low<br>high | **0.86 [0.79-0.92]**<br>1.29 [1.19-1.38] | 1.12 [1.06-1.20]<br>1.08 [0.98-1.16] |
| MAP <= 60 mmHg instead of 65 | low<br>high | 1.11 [1.03-1.19]<br>1.23 [1.15-1.31] | 1.11 [1.04-1.19]<br>1.25 [1.16-1.34] |
| Raw lactate >= 2 mmol/L (Exclusion when missing) | low<br>high | 1.20 [1.06-1.35]<br>1.26 [1.16-1.40] | 1.29 [1.19-1.39]<br>1.20 [1.10-1.30] |
| Adjustment for auxilliary demographic variables | low<br>high | 1.54 [1.45-1.64]<br>1.16 [1.08-1.23] | 1.13 [1.07-1.21]<br>1.15 [1.08-1.23] |
| Logistic Regression as estimator | low<br>high | 1.09 [0.99-1.21]<br>1.24 [1.16-1.34] | 1.07 [1.01-1.13]<br>1.06 [0.99-1.13] |
| Excluding infrequent treatments (<5) | low<br>high | 1.38 [1.29-1.47]<br>1.17 [1.10-1.25] | 0.94 [0.88-1.00]<br>1.12 [1.04-1.19] |
| Excluding patients with missing treatments | low<br>high | 1.77 [1.43-2.00]<br>1.35 [1.18-1.47] | 1.15 [1.02-1.30]<br>0.95 [0.84-1.05] |
| Backdoors measured before treatments | low<br>high | 1.91 [1.68-2.17]<br>0.97 [0.86-1.09] | 1.10 [1.01-1.22]<br>1.02 [0.92-1.14] |
| Limiting MIMIC to eICU collection time period (2014-2016) | low<br>high | 1.27 [1.19-1.36]<br>1.19 [1.12-1.27] | 1.12 [1.06-1.20]<br>1.02 [0.96-1.09] |

Table S10: Quantile odds between dose excesses and lack of clinical improvement in the eICU for different selection criteria after stratifying for severity (SOFA). Odds are presented as median and 95% CI. Odds significantly lower than 1 are reported in bold.

##### *B.4. Study Design and ML Pipeline*

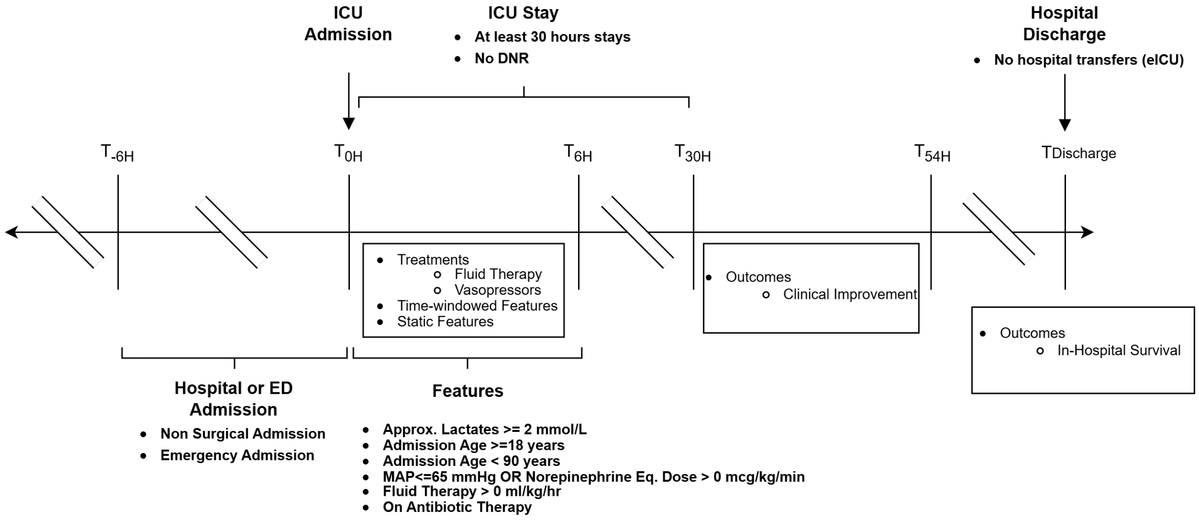


Figure S1: Study Design.

Alt text: Timeline of the study design. ICU admission is at hour zero, preceded by the earliest hospital interaction six hours earlier.

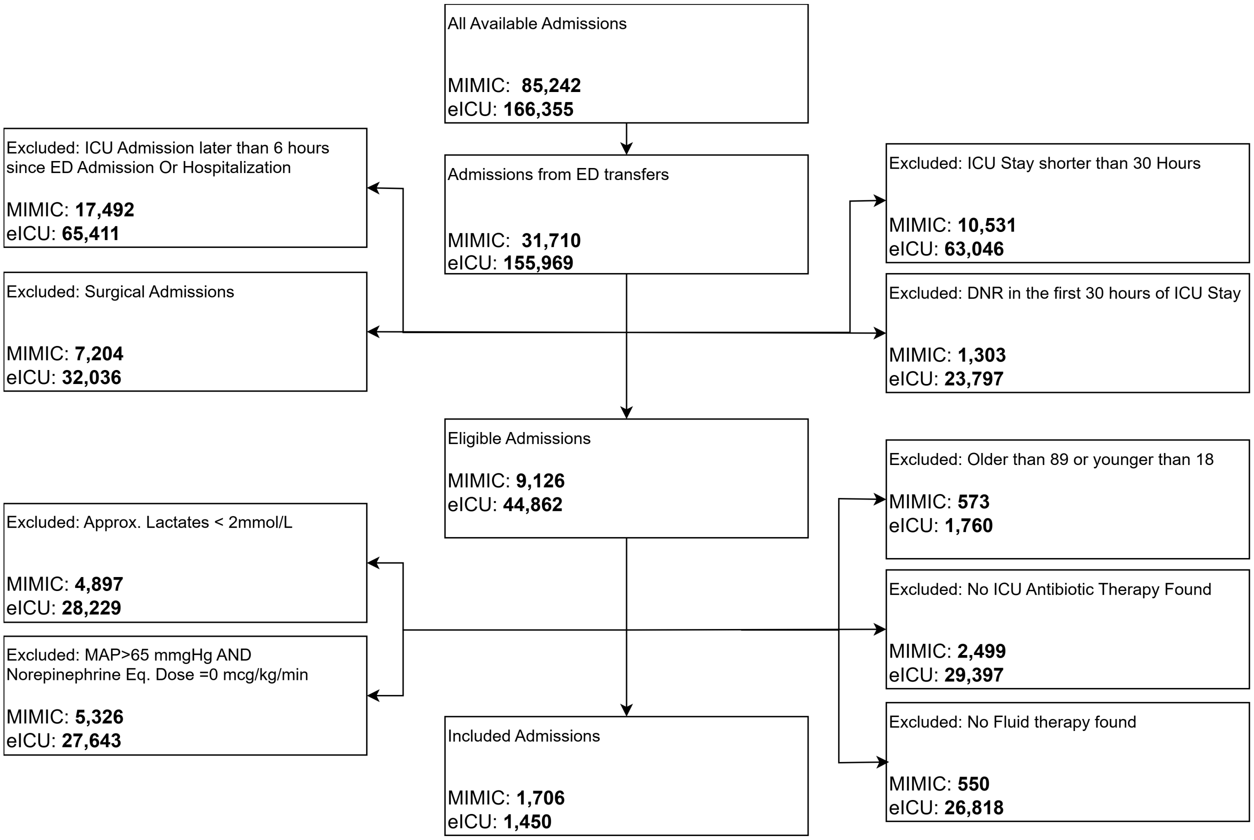


Figure S2: Inclusion and exclusion criteria. Criteria were assessed in parallel rather than sequentially, so a single admission may fail several of them; the per-criterion counts are therefore not mutually exclusive and do not sum to the number of admissions removed.

Alt text: Flow diagram of cohort selection for MIMIC and eICU, showing the starting number of admissions, the count removed by each exclusion criterion, and the final analysed cohorts of 1,706 and 1,450 admissions.

#### C. Causal Inference Details

##### C.1. Methodological details

Causal inference enables ML models to move beyond correlation and differentiate causation from mere statistical association. This differentiation is guided by a set of assumptions derived from domain knowledge, which in our case draws on existing evidence about sepsis and ICU dynamics. While many established causal frameworks exist, each encoding the causal assumptions differently, our framework of choice are Structural Causal Models (SCMs). SCMs are causal graphical model where causal assumptions are represented as directed arrows in a directed acyclic graph (DAG). This graphical representation acts as a navigational map governed by the strong theoretical foundations of the do-calculus, which determines the possibility of robustly quantifying interventional "what-if" simulation scenarios. Beyond their theoretical grounding, the causal graph offers a natural, visually intuitive language that fosters multidisciplinarity by enabling healthcare professionals, STEM practitioners, and data scientists to reason using a shared, common representation. In our case, the resulting causal graph, is shown in Fig. 2, and the clinical "what-if" question we aim to quantify is:

"What is the probability of survival (Y) if we administer a dosage of norepinephrine (N) and intravenous fluid (F) in the early hours following ICU admission in septic shock patients sharing clinical baseline (X)?"

This can be formalized as a *causal contrast*, here the Conditional Average Treatment Effect (CATE), as

$$\begin{aligned}&\text{CATE}((n,f),\mathbf{X}) =\\ &P(Y|do(N=n),do(F=f),\mathbf{X}) - P(Y|do(N=0),do(F=0),\mathbf{X}),\end{aligned}$$

where $do(N=n)$ and $do(F=f)$ represent *do* operators, corresponding to the act of intervening on a set of variables, in our case intravenous fluid dose $F$ and norepinephrine $N$, forcing them to take specific values $n$ and $f$ respectively, regardless of their causal parents. Accordingly, $P(Y|do(N=0),do(F=0),\mathbf{X})$ represents the probability of outcome $Y$ (where $Y=1$ in our case is either in-hospital survival or clinical improvement as opposed to $Y=0$) following intervention on $F$ and $N$ in a population with baseline clinical profile $\mathbf{X}$, equivalent in principle to performing an RCT. The CATE therefore represents the change in the probability of in-hospital survival or clinical improvement when delivering a combination of norepinephrine and fluids compared to no treatment. We employed the *Backdoor Criterion* to identify the features to include in the model. The Backdoor criterion can be verified from the causal graph alone and states that by adjusting for all parents $\mathbf{Z}$ of $F$ and $N$, which correspond to a subset of $\mathbf{X}$, the causal query of interest can be approximated from the data[9](for a rigorous formulation of the Backdoor definition, please refer to corollary 3 in[2]). In our case, as shown in Fig. S3, the criterion is satisfied and the CATE can be estimated by using

$$\begin{aligned}&P(Y|do(N=n),do(F=f),\mathbf{W}) =\\ &\sum_{\mathbf{z}} P(Y|N=n,F=f,\mathbf{Z}=\mathbf{z},\mathbf{W})P(\mathbf{Z}=\mathbf{z}|\mathbf{W}),\end{aligned}$$

where $\mathbf{W}$ are the remaining clinical descriptors in $\mathbf{X}$ that are not in $\mathbf{Z}$ and are non-descendants of $N$ and $F$, as in the main text.

The CATE can then be estimated in many different ways.[6,5,1,8] In our study, we adopt gradient boosting trees, an approach equivalent to an S-learner in meta-learner terminology[6]). A formal graphical representation of the causal assumptions among $\mathbf{Z}$, $\mathbf{X}$, $\mathbf{W}$, $N$ and $F$ is presented in Fig. S3 in B.4.

##### *C.2. Causal Graph Derivation Commentary*

To derive the causal graph represented in Fig. 2, we focused primarily on hemodynamic stability and the role of intravenous fluids administration and vasopressors dosage in the early hours of ICU. While we acknowledge that this constitutes a simplification of reality, it provides a structural rationale grounded in the clinical decision process. The graph incorporates both observed variables, those we retrieved from both the MIMIC and eICU datasets, and unmeasured ones, which either we could not reliably extract or were not available in the Electronic Health Records (EHR) (shown in grey in Fig. 2).

The derivation of the causal graph was central to defining the blueprint of our causal model and represents a major contributing factor in the results we obtained. Consensus on the causal graph was built in collaboration with expert causal methodologists, data scientists, and physicians, iteratively verifying that the encoded assumptions were clinically sound, that variables could be estimated from the data, and that causal backdoors could be approximated from the current assumptions.

The graph presented in Fig. 2 can be broadly read from left to right in chronological order, starting from infections acquired before Emergency Department (ED) admission to early antibiotic choices up until early treatments decisions in the ICU. Starting from the patient's ICU admission, we assume that appropriate antibiotics and fluid intake therapy were administered during the ER stay, as no data are available to reliably estimate them. From this points, we trace the clinician's decision process: the primary target is to stabilize tissue perfusion, which corresponds, as per guidelines, to setting a hemodynamic target of 65 mmHg of mean blood pressure. This can be achieved by acting on two levers: increasing cardiac output with fluids and increasing vascular resistance with vasopressors. Treatment doses are decided based on the progression of organ failure: norepinephrine equivalents are titrated based on signals of contractility, heart rate, and diastolic blood pressure. Renal function (via creatinine and urine output) and tissue perfusion (via lactates and mean blood pressure) inform clinicians on proper fluid dosing. In both cases, the patient's frailty is accounted for through their age as a proxy. The patient's response to infection is additionally captured through temperature and WBC count, which does not carry causal relevance in our model but enables stratification by approximate infection state.

The predictive model, used to benchmark the causal model's predictive performance, was built by including a broader set of routinely collected clinical and administrative markers from the ICU, in addition to all measured variables included in the causal graph. A full list of predictors is presented in Table S1.

##### *C.3. Model Training Details*

Training was performed via doubly nested cross-validation, where the inner loop iteratively sampled hyperparameters while the outer loop identified optimal calibration parameters. Model calibration was essential to represent model predictions as probabilities, which was necessary for estimating treatment effects. In the final outer scoring cross-validation loop, AUROCs and AUPRCs were estimated on the held-out outer folds, with AUROC selected as primary metric for its comparative properties.[7] In our experiment, for each outcome, we used 5 folds for the external scoring loop, 5 folds for the calibration loop, and 5 folds for the inner hyperparameter loop, each repeated for all 500 different hyperparameter combinations. AUROCs and AUPRCs in external validation were estimated over 2,000 bootstraps.

##### *C.4. Optimal Intervention Estimation and Validation Details*

The optimal treatment policy, defined as the combination of fluid and norepinephrine equivalent doses yielding the highest estimated treatment effect for a given patient, was estimated as

$$n^*, f^* = \underset{n,f}{\operatorname{argmax}} \quad \text{CATE}((n, f), \mathbf{X}),$$

where the optimization was performed across a discrete mesh grid of doses. The mesh grid's lower and upper limits were defined by the 5th and 95th percentiles observed in the training dataset. For each intervenable variable, i.e., norepinephrine and intravenous fluids, we examined 32 equidistant values in the mesh, equaling 1,024 different treatment combinations.

To validate the estimated policies, we compared patient outcomes in the external validation dataset according to the deviation between the administered treatment and the one suggested by the causal model. The rationale is that an informative policy should yield better outcomes for observed treatments aligning with model suggestions and worse prognoses for those diverging from them. To quantify this, we estimated the odds of worse outcomes for every 1-SD increase in the quantile-normalized deviation of the norepinephrine equivalent dose and of fluid intake, computed as the average of 1,000 bootstraps over 10 repeated 10-fold cross-validations on the external validation set. As a visual aid, we computed the differences between the observed and optimal doses of both norepinephrine equivalent and fluid intake, for each patient in external validation. We then grouped patients by deviation magnitude and estimated the average prevalence of both outcomes within each group. Deviations were discretized in bins of 0.05 mcg/kg/min and 1 ml/kg/hr for norepinephrine and fluid intake, respectively. Outcome prevalence within each bin was estimated over 1,000 bootstraps. To account for aleatoric uncertainty arising from documentation and aggregation biases in the measured interventions, we introduced a normal noise term with a standard deviation equal to the bin size. This procedure increases variance but better reflects our confidence in the quality of the documentation process. Finally, to assess heterogeneity of the treatment effect across varying patient baselines, we estimated treatment effects conditioned on covariates defined across a grid of the input space. We then broadly clustered profiles by common treatment-response trends to identify common trends. The most clinically informative examples were selected to illustrate the heterogeneous effects of vasopressors and fluid intake on in-hospital survival and clinical improvement.

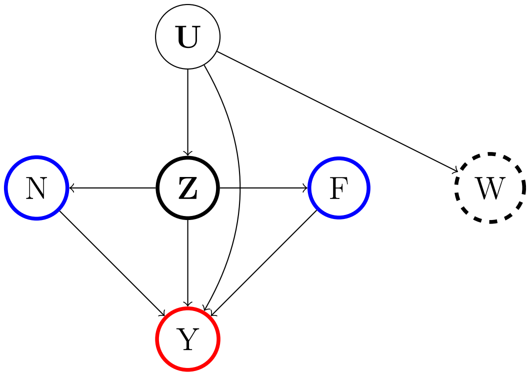


Figure S3: Formal causal assumptions for our study. Treatments $N$, $F$ are represented in bold blue, backdoors $\mathbf{Z}$ in bold black and the non-descendant conditioning descriptors $\mathbf{W}$ in dashed black, outcomes $Y$ in bold red.

Alt text: Small directed acyclic graph showing the formal causal assumptions: unmeasured variables U point to the backdoor set Z, to W and to the outcome Y; Z points to both treatments N and F and to Y; and both treatments point to Y.

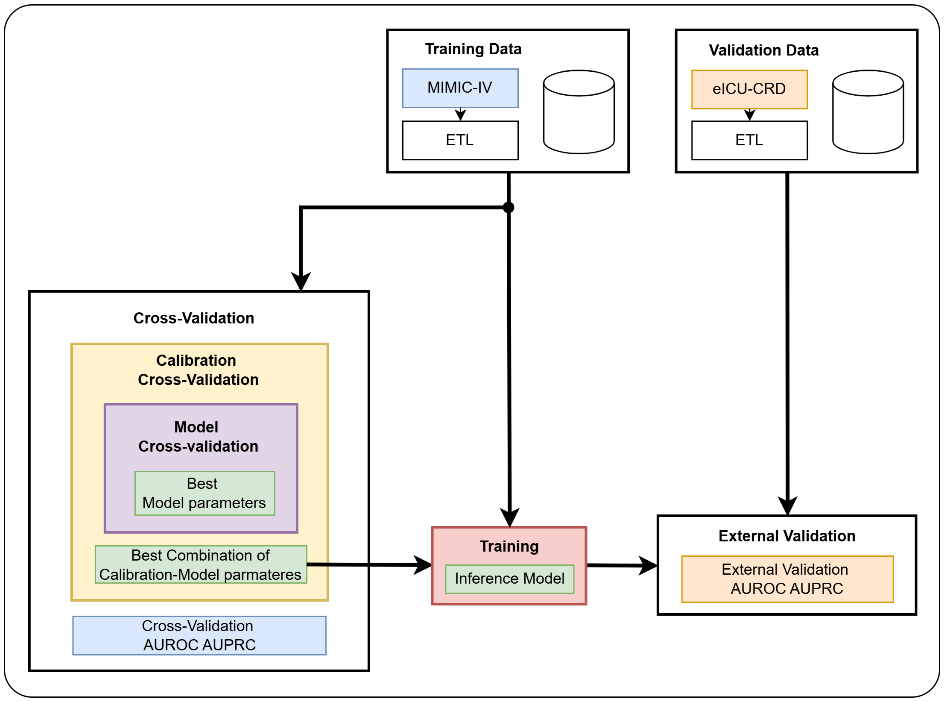


(a)

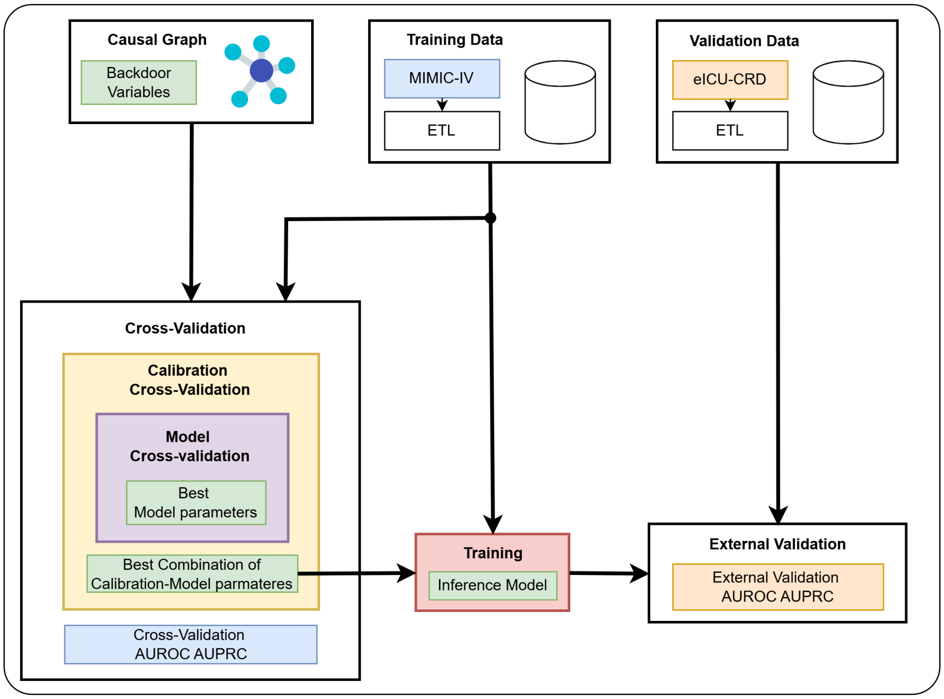


(b)

Figure S4: Training pipelines for the (S4a) Predictive model and (S4b) Causal model
Alt text: Two flow diagrams of the training pipelines, one for the predictive model and one for the causal model, each running from feature extraction through nested cross-validation and calibration to the fitted estimator.

| Feature Name | s14 | s20 | s38 | s49 | s55 | s56 | s96 | s118 | s130 | s143 | s157 | s167 |
|---|---|---|---|---|---|---|---|---|---|---|---|---|
| WBC [K/uL] | 18.1 | 12.1 | 10.3 | 18.1 | 26.0 | 12.1 | 18.1 | 16.1 | 12.1 | 18.1 | 26.0 | 26.0 |
| Residual SOFA | 1.8 | 2.8 | 0.0 | 2.0 | 3.8 | 3.8 | 2.0 | 2.0 | 0.8 | 2.0 | 1.8 | 2.8 |
| Temperature [C] | 37.0 | 36.4 | 36.9 | 37.2 | 37.6 | 36.5 | 37.2 | 35.9 | 35.9 | 37.2 | 36.7 | 37.2 |
| Urine Output [ml/kg/hr] | 1.7 | 0.9 | 2.4 | 0.5 | 0.5 | 0.1 | 0.5 | 0.9 | 1.2 | 0.5 | 0.4 | 0.7 |
| Mean Blood Pressure [mmHg] | 64.7 | 73.0 | 63.0 | 57.0 | 57.0 | 73.0 | 57.0 | 61.3 | 57.0 | 57.0 | 59.3 | 68.0 |
| Lactate [mmol/L] | 2.2 | 3.8 | 2.2 | 4.5 | 5.4 | 5.4 | 4.5 | 3.8 | 6.6 | 4.5 | 3.4 | 3.4 |
| Admission Age [Year(s)] | 74.0 | 49.0 | 49.0 | 70.0 | 74.0 | 49.0 | 70.0 | 84.0 | 66.0 | 70.0 | 84.0 | 66.0 |
| Diastolic Blood Pressure [mmHg] | 44.0 | 49.0 | 44.0 | 44.0 | 49.0 | 49.0 | 44.0 | 51.0 | 44.0 | 44.0 | 44.0 | 55.0 |
| Creatinine [mg/dL] | 1.1 | 1.0 | 1.4 | 2.1 | 4.0 | 2.7 | 2.1 | 1.4 | 1.4 | 2.1 | 1.3 | 4.0 |
| Heart Rate [BPM] | 85.3 | 75.0 | 97.1 | 75.0 | 91.5 | 97.1 | 75.0 | 91.5 | 109.7 | 75.0 | 109.7 | 80.6 |

Table S11: Clinical profiles of simulated cohorts.

### D. Reporting Guidelines

#### D.1. TRIPOD+AI

| Item No. | Item | Section(s) |
|---|---|---|
| 1 | Identify the study as developing or evaluating the performance of a multivariable prediction model, the target population, and the outcome to be predicted | Introduction |
| 2 | See TRIPOD+AI for Abstracts checklist | |
| 3a | Explain the healthcare context (including whether diagnostic or prognostic) and rationale for developing or evaluating the prediction model, including references to existing models | Introduction |
| 3b | Describe the target population and the intended purpose of the prediction model in the context of the care pathway, including its intended users (e.g., healthcare professionals, patients, public) | Introduction |
| 3c | Describe any known health inequalities between sociodemographic groups | Methods: Model Training and Predictive Performance Assessment |
| 4 | Specify the study objectives, including whether the study describes the development or validation of a prediction model (or both) | Introduction; Methods: Assumptions and Causal Implications |
| 5a | Describe the sources of data separately for the development and evaluation datasets (e.g., randomised trial, cohort, routine care or registry data), the rationale for using these data, and representativeness of the data | Methods: Cohort Derivation |
| 5b | Specify the dates of the collected participant data, including start and end of participant accrual; and, if applicable, end of follow-up | Methods: Cohort Derivation |
| 6a | Specify key elements of the study setting (e.g., primary care, secondary care, general population) including the number and location of centres | Methods: Cohort Derivation |
| 6b | Describe the eligibility criteria for study participants | Methods: Cohort Derivation |
| 6c | Give details of any treatments received, and how they were handled during model development or evaluation, if relevant | Methods: Cohort Derivation; Methods: Model Training and Predictive Performance Assessment |
| 7 | Describe any data pre-processing and quality checking, including whether this was similar across relevant sociodemographic groups | Methods: Cohort Derivation |
| 8a | Clearly define the outcome that is being predicted and the time horizon, including how and when assessed, the rationale for choosing this outcome, and whether the method of outcome assessment is consistent across sociodemographic groups | Methods: Cohort Derivation |
| 8b | If outcome assessment requires subjective interpretation, describe the qualifications and demographic characteristics of the outcome assessors | |
| 8c | Report any actions to blind assessment of the outcome to be predicted | |
| 9a | Describe the choice of initial predictors (e.g., literature, previous models, all available predictors) and any pre-selection of predictors before model building | Methods: Causal Graph Derivation |
| 9b | Clearly define all predictors, including how and when they were measured (and any actions to blind assessment of predictors for the outcome and other predictors) | Methods: Cohort Derivation; Supplementary B |
| 9c | If predictor measurement requires subjective interpretation, describe the qualifications and demographic characteristics of the predictor assessors | |
| 10 | Explain how the study size was arrived at (separately for development and evaluation), and justify that the study size was sufficient to answer the research question. Include details of any sample size calculation | Methods: Cohort Derivation |
| 11 | Describe how missing data were handled. Provide reasons for omitting any data | Methods: Cohort Derivation; Methods: Model Training and Predictive Performance Assessment; Supplementary B |
| 12a | Describe how the data were used (e.g., for development and evaluation of model performance) in the analysis, including whether the data were partitioned, considering any sample size requirements | Methods: Model Training and Predictive Performance Assessment |
| 12b | Depending on the type of model, describe how predictors were handled in the analyses (functional form, rescaling, transformation, or any standardisation). | Methods: Cohort Derivation; Methods: Model Training and Predictive Performance Assessment |
| 12c | Specify the type of model, rationale, all model-building steps, including any hyperparameter tuning, and method for internal validation | Methods: Model Training and Predictive Performance Assessment |

| Item No. | Item | Section(s) |
|---|---|---|
| 12d | Describe if and how any heterogeneity in estimates of model parameter values and model performance was handled and quantified across clusters (e.g., hospitals, countries). See TRIPOD-Cluster for additional considerations | |
| 12e | Specify all measures and plots used (and their rationale) to evaluate model performance (e.g., discrimination, calibration, clinical utility) and, if relevant, to compare multiple models | Methods: Model Training and Predictive Performance Assessment |
| 12f | Describe any model updating (e.g., recalibration) arising from the model evaluation, either overall or for particular sociodemographic groups or settings | |
| 12g | For model evaluation, describe how the model predictions were calculated (e.g., formula, code, object, application programming interface) | Methods: Model Training and Predictive Performance Assessment; Methods: Optimal Intervention Estimation and Validation |
| 13 | If class imbalance methods were used, state why and how this was done, and any subsequent methods to recalibrate the model or the model predictions | |
| 14 | Describe any approaches that were used to address model fairness and their rationale | Methods: Model Training and Predictive Performance Assessment |
| 15 | Specify the output of the prediction model (e.g., probabilities, classification). Provide details and rationale for any classification and how the thresholds were identified | Methods: Optimal Intervention Estimation and Validation |
| 16 | Identify any differences between the development and evaluation data in healthcare setting, eligibility criteria, outcome, and predictors | Results: Study Design and Cohort Characterization; Supplementary B |
| 17 | Name the institutional research board or ethics committee that approved the study and describe the participant-informed consent or the ethics committee waiver of informed consent | Data Sharing Statement |
| 18a | Give the source of funding and the role of the funders for the present study | Funding |
| 18b | Declare any conflicts of interest and financial disclosures for all authors | Declaration of interests |
| 18c | Indicate where the study protocol can be accessed or state that a protocol was not prepared | |
| 18d | Provide registration information for the study, including register name and registration number, or state that the study was not registered | |
| 18e | Provide details of the availability of the study data | Data Sharing Statement |
| 18f | Provide details of the availability of the analytical code | Data Sharing Statement |
| 19 | Provide details of any patient and public involvement during the design, conduct, reporting, interpretation, or dissemination of the study or state no involvement. | |
| 20a | Describe the flow of participants through the study, including the number of participants with and without the outcome and, if applicable, a summary of the follow-up time. A diagram may be helpful. | Results: Study Design and Cohort Characterization; Supplementary B.4 |
| 20b | Report the characteristics overall and, where applicable, for each data source or setting, including the key dates, key predictors (including demographics), treatments received, sample size, number of outcome events, follow-up time, and amount of missing data. A table may be helpful. Report any differences across key demographic groups. | Results: Study Design and Cohort Characterization; Supplementary B |
| 20c | For model evaluation, show a comparison with the development data of the distribution of important predictors (demographics, predictors, and outcome). | Supplementary B |
| 21 | Specify the number of participants and outcome events in each analysis (e.g., for model development, hyperparameter tuning, model evaluation) | Methods: Model Training and Predictive Performance Assessment; Results: Study Design and Cohort Characterization; Supplementary B.2 |
| 22 | Provide details of the full prediction model (e.g., formula, code, object, application programming interface) to allow predictions in new individuals and to enable third-party evaluation and implementation, including any restrictions to access or re-use (e.g., freely available, proprietary) | Data Sharing Statement |
| 23a | Report model performance estimates with confidence intervals, including for any key subgroups (e.g., sociodemographic). Consider plots to aid presentation. | Results: Optimal treatments, confounding and clinical outcomes; Supplementary B.2 |
| 23b | If examined, report results of any heterogeneity in model performance across clusters. See TRIPOD Cluster for additional details. | Results: Heterogeneity of the treatment effect |
| 24 | Report the results from any model updating, including the updated model and subsequent performance | |

| Item No. | Item | Section(s) |
|---|---|---|
| 25 | Give an overall interpretation of the main results, including issues of fairness in the context of the objectives and previous studies | Discussion |
| 26 | Discuss any limitations of the study (such as a non-representative sample, sample size, overfitting, missing data) and their effects on any biases, statistical uncertainty, and generalizability | Discussion |
| 27a | Describe how poor quality or unavailable input data (e.g., predictor values) should be assessed and handled when implementing the prediction model | Discussion |
| 27b | Specify whether users will be required to interact in the handling of the input data or use of the model, and what level of expertise is required of users | |
| 27c | Discuss any next steps for future research, with a specific view to applicability and generalizability of the model | Discussion |

Table S12: TRIPOD+AI Checklist[4]

##### *D.2. TARGET Statement*

| Item No. | Item | Section(s) |
|---|---|---|
| 1a | Identify that the study attempts to emulate a target trial using observational data. State the study objectives and briefly summarize the specified target trial. | Introduction |
| 1b | Report the data sources used for emulation. | Introduction |
| 1c | Summarize key assumptions, statistical methods, findings and conclusions. | Introduction |
| 2 | Describe the scientific background of the study and the gap in knowledge. | Introduction |
| 3 | Summarize the causal question. | Methods: Assumptions and Causal Implications; Results: Causal graph derivation and physiological grounding |
| 4 | Describe the rationale for emulating a target trial with the available data. Cite randomized trials informing the design of the target trial if applicable. | Introduction |
| 5 | Cite the data sources contributing to the analyses and for each one describe the following: original purpose, type, the geographic locations, setting and time-period. If relevant, describe how the data were linked or pooled. | Methods: Cohort Derivation |
| 6a | Target trial specification: Describe the eligibility criteria. | Methods: Cohort Derivation; Results: Study Design and Cohort Characterization |
| 7a | Target trial emulation: Describe how the eligibility criteria were operationalized with the data. | Methods: Cohort Derivation; Supplementary B |
| 6b | Target trial specification: Describe the treatment strategies that would be compared. | Methods: Causal Graph Derivation; Results: Study Design and Cohort Characterization |
| 7b | Target trial emulation: Describe how the treatment strategies were operationalized with the data. | Methods: Cohort Derivation; Results: Study Design and Cohort Characterization; Supplementary B |
| 6c | Target trial specification: Report that eligible individuals would be randomly assigned to treatment strategies and may be aware of their treatment allocation. | Methods: Assumptions and Causal Implications; Results: Optimal treatments, confounding and clinical outcomes |
| 7c | Target trial emulation: Describe how assignment to treatment strategies was operationalized with the data. | Methods: Assumptions and Causal Implications; Methods: Optimal Intervention Estimation and Validation; Results: Optimal treatments, confounding and clinical outcomes |
| 6d | Target trial specification: Clarify that follow-up would start at time of assignment to the treatment strategies. Specify when follow-up would end. | Methods: Cohort Derivation; Results: Study Design and Cohort Characterization |

| Item No. | Item | Section(s) |
|---|---|---|
| 7d | Target trial emulation: Clarify that follow-up starts at the time individuals were assigned to the treatment strategies. Describe how the end of follow-up was operationalized with the data. | Methods: Cohort Derivation |
| 6e | Target trial specification: Describe the outcomes. | Methods: Cohort Derivation; Results: Study Design and Cohort Characterization |
| 7e | Target trial emulation: Describe how the outcomes were operationalized with the data. | Methods: Cohort Derivation; Supplementary B |
| 6f | Target trial specification: Describe the causal contrasts of interest, including effect measures. | Methods: Assumptions and Causal Implications; Results: Optimal treatments, confounding and clinical outcomes |
| 7f | Target trial emulation: Describe how the causal contrasts were operationalized with the data, including effect measures. | Methods: Model Training and Predictive Performance Assessment; Methods: Optimal Intervention Estimation and Validation; Results: Optimal treatments, confounding and clinical outcomes |
| 6g | Target trial specification: Describe assumptions that would be made to identify each causal estimand. Describe the variables, if any, related to these assumptions. | Methods: Assumptions and Causal Implications; Methods: Causal Graph Derivation; Results: Optimal treatments, confounding and clinical outcomes |
| 7g.i | Target trial emulation: For each causal estimand, describe assumptions made to identify it, including assumptions regarding baseline confounding due to lack of randomization. | Methods: Assumptions and Causal Implications; Methods: Causal Graph Derivation; Results: Optimal treatments, confounding and clinical outcomes |
| 7g.ii | Target trial emulation: Describe how the variables related to these assumptions were operationalized with the data. | Methods: Cohort Derivation; Supplementary B |
| 6h | Target trial specification: For each causal estimand, describe the data analysis procedures and any associated statistical modelling assumptions, including approaches for handling missing data. | Methods: Model Training and Predictive Performance Assessment; Methods: Optimal Intervention Estimation and Validation |
| 7h.i | Target trial emulation: For each causal estimand, describe the data analysis procedures and any associated statistical modelling assumptions, including approaches for handling missing data. | Methods: Model Training and Predictive Performance Assessment; Methods: Optimal Intervention Estimation and Validation |
| 7h.ii | Target trial emulation: For each causal estimand, describe any additional analyses conducted to assess the sensitivity of the results to the choice of operationalizations, assumptions and analysis. | Methods: Sensitivity Analysis; Results: Robustness to design choices; Supplementary B.3 |
| 8 | Report numbers of individuals assessed for eligibility, eligible, and assigned to each treatment strategy. A flow diagram is strongly recommended. | Results: Study Design and Cohort Characterization; Supplementary B |
| 9 | Describe the distribution of characteristics of individuals at baseline, by treatment strategy. | Results: Study Design and Cohort Characterization; Supplementary B |
| 10 | Summarize length of follow-up and describe reasons for end of follow-up for each treatment strategy and causal contrast. | Methods: Cohort Derivation; Results: Study Design and Cohort Characterization |

| Item No. | Item | Section(s) |
|---|---|---|
| 11 | Describe the frequency of missing data in all variables, by treatment strategy when applicable. | Supplementary B |
| 12 | Describe the frequency or distribution of each outcome, by treatment strategy. | Results: Study Design and Cohort Characterization; Supplementary B |
| 13 | Report the effect estimates for each causal contrast with corresponding measures of precision, including both absolute and relative measures of effect, when applicable. | Results: Optimal treatments, confounding and clinical outcomes; Results: Heterogeneity of the treatment effect; Supplementary B.3 |
| 14 | Report results of all analyses to assess the sensitivity of the estimates to choices in operationalizations, assumptions and analysis. | Results: Robustness to design choices; Supplementary B.3 |
| 15 | Provide an interpretation of the key findings. | Discussion |
| 16 | Discuss the limitations of the study considering differences between the target trial and its emulation and the plausibility of assumptions, including assumptions regarding baseline confounding due to lack of randomization. | Discussion |
| 17 | Provide the institutional research board or ethics committee that approved the study and approval numbers, if relevant. | Data Sharing Statement |
| 18 | State whether, when and where the study protocol was registered. | |
| 19 | Provide information on whether data, analytic code and/or other materials are accessible, and where and how they can be accessed. | Data Sharing Statement |
| 20 | Provide the sources of funding and detail the role of the funders in the design, conduct and reporting of the study. | Funding |
| 21 | State any conflicts of interest and financial disclosures for all authors. | Declaration of interests |

Table S13: Target Statement Checklist[3]
